# Tree-informed Bayesian multi-source domain adaptation: cross-population probabilistic cause-of-death assignment using verbal autopsy

**DOI:** 10.1101/2021.12.20.21268145

**Authors:** Zhenke Wu, Zehang R. Li, Irena Chen, Mengbing Li

## Abstract

Determining causes of deaths (COD) occurred outside of civil registration and vital statistics systems is challenging. A technique called verbal autopsy (VA) is widely adopted to gather information on deaths in practice. A VA consists of interviewing relatives of a deceased person about symptoms of the deceased in the period leading to the death, often resulting in multivariate binary responses. While statistical methods have been devised for estimating the cause-specific mortality fractions (CSMFs) for a study population, continued expansion of VA to new populations (or “domains”) necessitates approaches that recognize between-domain differences while capitalizing on potential similarities. In this paper, we propose such a domain-adaptive method that integrates external between-domain similarity information encoded by a pre-specified rooted weighted tree. Given a cause, we use latent class models to characterize the conditional distributions of the responses that may vary by domain. We specify a logistic stick-breaking Gaussian diffusion process prior along the tree for class mixing weights with node-specific spike-and-slab priors to pool information between the domains in a data-driven way. Posterior inference is conducted via a scalable variational Bayes algorithm. Simulation studies show that the domain adaptation enabled by the proposed method improves CSMF estimation and individual COD assignment. We also illustrate and evaluate the method using a validation data set. The paper concludes with a discussion on limitations and future directions.

## 1. Introduction

### 1.1 Verbal Autopsy (VA): background

Patterns of mortality and causes of death at the community level are critical to informing public health policies, tracking trends, and prioritizing interventions for local governments and public health officials. Civil registration systems that track births, deaths and their causes provide the basis for countries to identify their most pressing health issues. Despite ongoing global efforts to strengthen the civil registration and vital statistics (CRVS) system, two-thirds of 56 million annual deaths go unrecorded, especially in low- and middle-income countries (LMIC), leaving glaring gaps for reliable mortality information (World Health Organization, 2021). Verbal autopsy (VA) is one of the most well-established and realistic methods to collect information about cause of death (COD) in these situations when medically certified cause of death is unavailable. VAs collect information on deaths by interviewing caregivers (or individuals who witnessed the death) of the deceased. Typically, information about healthcare access, demographic information, and various indicators of symptoms leading to the death are collected. See Chandramohan *and others* (2021) for a recent review of historical developments, ongoing efforts to standardize VA instruments and implications for LMIC.

The central analytic goal is to use VA data to derive population-level cause-specific mortality fractions (CSMFs) and to produce individual-level COD assignment. In particular, VA data contain pertinent information on signs, symptoms, and circumstances leading to death, generically referred to as “symptoms” in this paper. These symptoms are often coded into binary “Yes”/”No” answers, resulting in data sets that contain multiple binary responses for each death. Algorithmic and probablistic methods have been developed to automate the task of estimating CSMFs and individual-level COD assignment. See Li *and others* (2021*b*) for an excellent comprehensive review of the major methods and software as well as the references therein for recent methodological improvements. Developments in analytic methods and reproducible open-source software for VA have greatly fostered confidence in large-scale implementations of VA in many LMICs.

#### 1.1.1 Statistical challenges in domain adaptation in VA research

However, one emerging analytical challenge in expanding VA to new populations is in developing approaches that recognize and address potential differences between the existing and new populations in terms of the joint distribution of causes of death and VA responses. Such a problem is an example of “domain adaptation” in machine learning literature (e.g., Pan and Yang, 2009), but has received relatively little attention in VA research. In particular, CSMFs comprise a vector of population-level marginal probabilities of the causes and may differ by domain; this is most natural because a cause may differentially contribute to deaths occurred in different study populations. The conditional distribution of the VA responses given a cause characterizes symptom-cause relationships and may also differ by domain; we focus our paper and model formulation on addressing this aspect of domain adaptation.

It is well known that more accurate estimation of the conditional distribution of the multivariate binary VA responses given a cause can result in substantial improvements to CSMF estimation performance (e.g., Kunihama *and others*, 2020; Li, McComick and Clark, 2020). To acknowledge potential between-domain differences in these conditional distributions given a cause, it is therefore tempting to directly estimate them for each domain separately. However, in a domain with few sampled deaths due to a cause, such direct estimates are often vulnerable to statistical instability, hindering accurate CSMF estimation. This issue worsens still if for that same cause the numbers of sampled deaths are small in multiple domains. In such cases, pooling information from similar domains would improve the estimation of these conditional distributions which in turn would propagate to improving the estimation of CSMFs. On the other hand, an extreme complete-pooling approach that forces domains to have an identical conditional distribution of the VA symptoms given any cause is restrictive and would obscure the study of important between-domain variations in response patterns (e.g., King and Lu, 2008; McCormick *and others*, 2016). Data-driven pooling of information between the domains for each cause is desirable.

#### 1.1.2 Existing literature

Here we briefly describe a few existing work related to domain adaptation in verbal autopsy studies and how the proposed method differ from them. Datta *and others* (2021) and Fiksel *and others* (2021) developed methods that calibrate CSMF estimates obtained from VA algorithms trained on a training data set to produce CSMF estimates in a new population. These calibration methods differ from our work in three important ways. First, such calibration methods only consider the estimated CSMFs from a list of trained VA algorithms, and are hence not designed for using individual-level information in the training data set to perform calibration. Second, the calibration relies on a small number of deaths with medically-confirmed causes in the new population. Third, causes often need to be manually combined before calibration can be applied to produce stable and meaningful results. Our work focus on using all individual-level data from multiple populations (referred to as “source domains”) with known causes of death, and cause-of-death assignment in a new population (referred to as “target domain”). The proposed method does not require known cause-of-death labels in the target domain or any ad hoc collapsing of causes.

Among a few methods that directly model the individual-level VA data under domain adaptation, one related work is Moran *and others* (2021) that introduces a factor regression method to let the conditional distribution of the VA symptoms given a cause vary by additional individual-level covariates, which may include dummy domain indicators. This approach again is not designed for the scenario where cause-of-death labels are fully unobserved in the target domain. Our work is most related to Li *and others* (2021*c*), where a latent class model framework was proposed to model the conditional dependence relationship among symptoms with improved interpretability and computational speed. However VAs collected from different domains are treated as marginally independent data sets. Our work significantly extends Li *and others* (2021*c*) by proposing a framework that can integrate additional external structural information across the domains. This allows us to efficiently pool information across domains and improve the stability of the estimates when some causes are rarely observed.

### 1.2 Main contributions

This paper develops a novel tree-integrative framework for CSMF estimation and individual-level COD assignment based on latent class models (Lazarsfeld, 1950) that jointly models multivariate binary data obtained from multiple source domains and a target domain. Our framework explicitly acknowledges domain-by-domain variation in the distribution of causes and distribution of symptoms given causes. Most importantly, it takes into account the structural similarities among deaths from related domains. Our main methodological contributions are two folds.

First, we propose a data-driven pooling of information across domains via a pre-specified hierarchy represented by a rooted weighted tree. This approach is shown to encourage similar conditional dependence structure across domains while recognizing important between-domain differences resulting in more accurate CSMF estimation. Simulation studies show the proposed approach has better performance compared to estimates that either completely, incorrectly, or minimally pool information across domains. Although the method is general, in this paper, we illustrate the method by a domain tree defined by geographical region of each study site, which serves as a proxy for potential regional variations in factors which may drive differences in the conditional distributions of symptoms given a cause, e.g., VA interviewer training, culture in symptom disclosure of a deceased, etc. As a secondary feature, the proposed approach also uses a hierarchy over the causes to enable information pooling between causes so that a rare cause can be pooled with similar causes to produce more stable estimates of the class-specific response profiles.

Second, we propose a tree-based measure of dissimilarity in symptom-cause relationship between the target domain and each of the source domains, separately for each cause. The proposed measure admits rich interpretation of empirical evidence about the manner in which causes differ in between-domain similarities. For example, causes with highly recognizable and specific symptoms (e.g., “Drowning”) may have symptom-cause conditional distributions that remain similar regardless of the domains, while less so for other causes with complex etiologies that are prone to differential reporting patterns across the domains.

#### Paper organization

The rest of the paper is organized as follows. Section 2 reviews tree-related terminologies and presents the proposed model. Section 3.1 specifies prior distributions. A variational Bayes algorithm is presented in Section 4. Section 5 conducts simulation studies to illustrate the operating characteristics of the proposed method. In Section 6, we use a validation data set to illustrate the method. The paper concludes with a brief summary and discussion on limitations and some future directions.

## 2. Model

We first introduce necessary terminologies and notations for characterizing a rooted weighted tree. The proposed nested latent class model (NLCM) is then formulated for deaths, each with an observed link to a leaf in a tree over source and target domains.

### 2.1 Rooted weighted trees

A rooted tree is a graph 𝒯 = (𝒱, *E*) with node set 𝒱 and edge set *E* where there is a root *u*_0_ and each node has at most one parent node. Let *p* = |𝒱| represent the total number of leaf and non-leaf nodes. Let 𝒱_leaf_ ⊂ 𝒱 be the set of leaves (i.e., nodes without children), and *p*_leaf_ = |𝒱_leaf_ | *< p*. We typically use *u* to denote any node (*u* ∈ 𝒱) and *v* to denote any leaf (*v* ∈ 𝒱_leaf_). Each edge in a rooted tree defines a *clade*: the group ofleaves below it. Splitting the tree at an edge creates a partition of the leaves into two groups. For any node *u* ∈ 𝒟, the following notations apply: *c*(*u*) is the set of offspring of *u, pa*(*u*) is the parent of *u, d*(*u*) is the set of descendants of *u* including *u*, and *a*(*u*) is the set of ancestors of *u* including *u*. At the top of Figure 1, a hypothetical tree for *G* = 4 source domains and one target domain with *p* = 8 and *p*_leaf_ = 5 is shown. If *u* = 2, then *c*(*u*) = {5, 6}, *pa*(*u*) = {1}, *d*(*u*) = {2, 5, 6}, and *a*(*u*) = {1, 2}. See Figure 3 (top margin) for an instance of a nested hierarchy for six domains where VA data are collected: *p*_leaf_ = 6 leaves representing six study sites, and *p* − *p*_leaf_ = 3 non-leaf nodes subsuming the six leaf descendants (root node representing “global”; two internal nodes representing two countries, “India” and “Tanzania”).

**Fig. 1:**
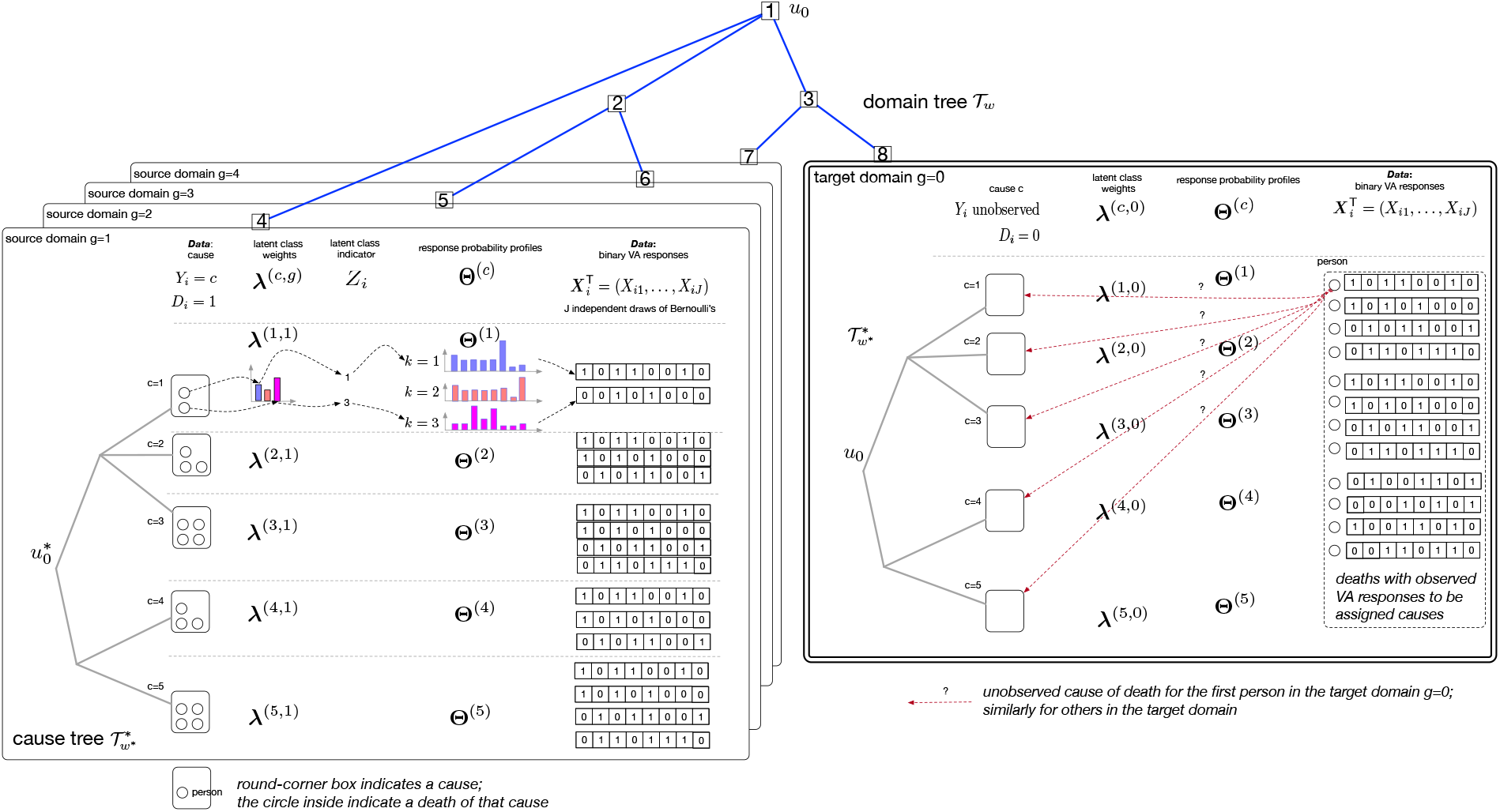
Schematic representation of the nested LCM model structure. *Top*) An eight-node (root *u* = *u*_0_) tree over five hypothetical domains is used to specify a tree-structured shrinkage prior for ***λ***^(*c,g*)^, *g* = 0, 1, …, *G*; *Left*) *G* = 4 source domains (*D*_*i*_ = 1, …, 4), shown in overlaid plates; causes of deaths are observed to be in one of *C* = 5 hypothetical causes. Hypothetical observed *J* = 8 binary VA responses are also shown; *Right*) one target domain where the causes of deaths *Y*_*i*_’s are unobserved but the binary VA responses ***X***_*i*_’s are observed. The cause hierarchy, as a secondary feature, is represented by a tree with five leaves representing *C* = 5 causes. Three latent classes (*K* = 3) are illustrated here.

Edge-weighted graphs appear as a model for numerous problems where nodes are linked with edges of different weights. In particular, the edges in 𝒯 are attached with weights where *w* : *E* → ℝ+ is a weight function. Let 𝒯_*w*_ = (𝒯, *w*) be a rooted weighted tree. A path in a graph is a sequence of edges which joins a sequence of distinct vertices. For a path *P* in the tree connecting two nodes, *w*(*P*) is defined as the sum of all the edge weights along the path, often referred to as the “length” of *P*. The distance between two vertices *u* and *u*, denoted by *dist*_T*w*_ (*u, u*) is the length of a shortest (with minimum length) (*u, u*)-path. *dist*_T*w*_ is a distance: it is symmetric and satisfies the triangle inequality. In this paper, we use *w*_*u*_ to represent the edge length between a node *u* and its parent node *pa*(*u*). *w*_*u*_ is fully determined by 𝒯_*w*_. For the root *u*_0_, there are no parents, i.e. *pa*(*u*_0_) = ∅; we set *w*_*u*0_ = 1. In VA contexts, although *w*_*u*_ may be specified via the dendrogram resulting from a hierarchical clustering of domain-level covariates, in Section 6 we will set *w*_*u*_ = 1 to use minimal external domain similarity information (geographical region) for simpler exposition.

### 2.2 Nested latent class models (Nested LCM)

Although LCMs work for multiple discrete responses of more than two levels (e.g., Lazarsfeld, 1950), in this paper, we present the model for multivariate binary responses for simpler exposition.

#### Notations

Let ***X***_*i*_ = (*X*_*i*1_, …, *X*_*iJ*_)^T^ ∈ {0, 1}^*J*^ be a vector of *J* binary responses for subject *i* ∈ [*N*] where *N* is the total number of subjects; here [*Q*] = {1, …, *Q*} generically represents positive integers no greater than a positive integer *Q*. Let (*Y*_*i*_, *D*_*i*_) represent (cause of death, domain), where *Y*_*i*_ takes its value from {1, …, *C*} indicating the cause of death among a total of *C* pre-specified causes; let ***Y*** = (*Y*_1_, …, *Y*_*N*_)^T^. *D*_*i*_ takes its value from {0, 1, …, *G*} indicating subject *i*’s domain membership: 0 for target domain, and 1 to *G* for *G* pre-specified source domains. Let ***D*** = (*D*_1_, …, *D*_*N*_)^T^. Throughout the paper, *D*_*i*_ is assumed to be observed for all subjects; *Y*_*i*_ is assumed to be observed for subjects in the source domain {*i* : *D*_*i*_ ≠ 0} but unobserved for subjects in the target domain {*i* : *D*_*i*_ = 0}. Let ***Y*** ^obs^ = {*Y*_*i*_ : *D*_*i*_ *≠* 0} and ***Y*** ^mis^ = {*Y*_*i*_ : *D*_*i*_ = 0}; we then have ***Y*** = (***Y*** ^obs^, ***Y*** ^mis^)^T^. Let **X** = (***X***_1_, …, ***X***_*N*_)^T^ be an *N* × *J* binary data matrix for all subjects. ***D*** maps every row of data **X** to a leaf in the tree for domains 𝒯_*w*_. Similarities between domains are then characterized by between-domain distances in 𝒯_*w*_. Finally, let 𝒟 = (**X, *Y*** ^obs^, ***D***) represent the data from all the domains. We use “pr(*A* | *B*)” to represent the conditional density of random variable(s) in *A* given *B*.

##### 2.2.1 Model formulation We assume the following model specifications for 𝒟

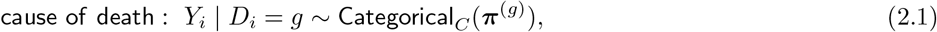

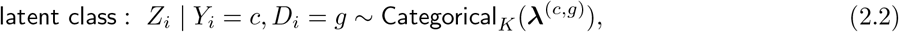

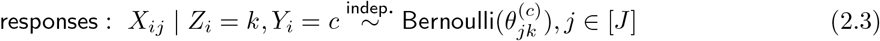

for *i* ∈ [*N*], *g* ∈ {0} ∪ [*G*], where the population parameters 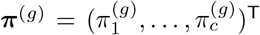 with 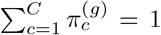 are referred to as “cause-specific mortality fractions” (CSMFs). Importantly, {***π***^(*g*)^, *g* = 0, 1, …, *G*} are not constrained to be identical. We seek to estimate ***π***^(0)^ and {*Y*_*i*_ : *D*_*i*_ = 0}.

On the latent classes, ***Z*** = (*Z*_1_, …, *Z*_*N*_)^T^ ∈ [*K*]^*N*^ is a vector of class memberships for all subjects; 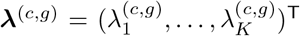 is a vector of class weights that sum to one: 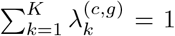. See Remark 2.1 for the nuisance role of ***Z*** and the nuisance notion of “class” that are introduced for inducing conditional stochastic dependence among VA responses given any pair of cause and domain. On terminology, by Equation (2.2), the *K* latent classes that *Z*_*i*_ can take are nested within a cause of death *c*; we refer to the model as “nested” latent class model.

On the class-specific response probabilities, for a subject died of cause 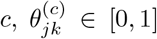 is the positive response probability for item *j* in class *k*. Let 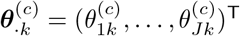 be the vector of the *k*-th class response probability profile; let 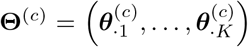 collect these probabilities into a *J* × *K* matrix with (*j, k*)-th element 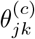. Note that **Θ**^(*c*)^ may vary by cause *c*. This admits flexible characterization of symptoms that may have distinct distributions for different true causes-of-death. In addition, for any given *c*, **Θ**^(*c*)^ is assumed domain-invariant (source or target) to facilitate shared interpretations. Conversely, letting the class response profiles vary greatly between domains would weaken the diagnostic explanability of the VA questionnaire items. See Figure 1 for a schematic representation of the data generating process under the proposed model.

For any given cause *c*, despite shared **Θ**^(*c*)^ across the domains, pr(***X***_*i*_ | *Y*_*i*_ = *c, D*_*i*_ = *g*) may differ by domain *g* as a result of distinct class weights ***λ***^(*c,g*)^ across the domains. This is readily seen from Equations (2.2) and (2.3) which imply that the conditional distributions are fully parameterized by (**Θ**^(*c*)^, ***λ***^(*c,g*)^):

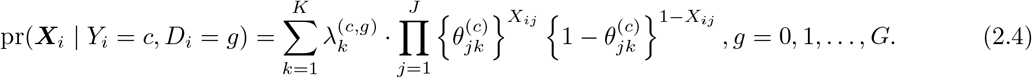

If ***λ***^*c,g*^ = ***λ***^(*c,g ′*^) for any *g, g ′* = 0, 1, …, *G*, Equation (2.4) simplifies to pr(***X***_*i*_ | *Y*_*i*_ = *c*).

Remark 2.1 The model treats ***Z*** and the notion of “class” as technical nuisances that are introduced for the sole purpose of flexibly modeling pr(***X***_*i*_ | *Y*_*i*_, *D*_*i*_) (Dunson and Xing, 2009). In particular, when *K ⩾* 2 and {Θ^(*c*)^} differ by *c*, although Equation (2.3) assumes conditional independence given a latent class and a cause, by integrating over *Z*_*i*_ with probabilities 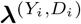, we induce stochastic dependence among the *J* components of ***X***_*i*_ (Equation (2.4)). Setting *K* = 1 would assume that the VA responses are mutually independent given any pair of cause and domain.

Remark 2.2 Equations (2.1) to (2.3) under *D*_*i*_ = 0 are equivalent to a *K* · *C*-class LCM for {***X***_*i*_ : *D*_*i*_ = 0} parameterized by (***π***^(0)^, ***λ***^(*c*,0)^, **Θ**^(*c*)^, *c* ∈ [*C*]). Fortunately, based on data from the source domains, the multivariate binary VA response data {***X***_*i*_ : *D*_*i*_ = 0} tabulated by the observed causes of deaths (***Y*** ^obs^) provide direct information for estimating **Θ**^(*c*)^ that is shared across the domains.

## 3. Priors

### 3.1 Tree-structured shrinkage prior

#### 3.1.1 Motivation and overview

Estimating target domain CSMFs and individual cause-of-death assignment rely on efficient learning of pr(***X***_*i*_ | *Y*_*i*_ = *c, D*_*i*_ = 0), the joint distribution of multivariate binary VA responses given each cause. In particular, by Bayes rule

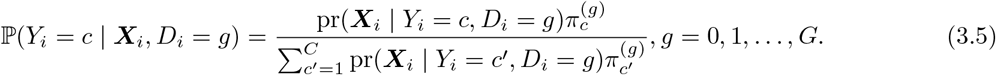

When *g* = 0, even with known ***π***^(0)^, a poor estimate of pr(***X***_*i*_ | *Y*_*i*_ = *c, D*_*i*_ = 0) can adversely impact the cause-of-death assignment on an individual level; when ***π***^(0)^ is unknown as in our context, a good estimate of the conditional distribution pr(***X***_*i*_ | *Y*_*i*_ = *c, D*_*i*_ = 0) remains critical to stable statistical estimation, e.g., via expectation-maximization for finite mixture models. The same issue persists for accurate individual-level cause-of-death assignment when *g* ≠ 0 for which ***π***^(*g*)^ can be directly estimated. However, obtaining a good estimate of pr(***X***_*i*_ | *Y*_*i*_ = *c, D*_*i*_ = *g*) is often challenging when there exist small or even zero cell counts for particular combinations of (*c, g*), which renders direct estimation of pr(***X***_*i*_ | *Y*_*i*_ = *c, D*_*i*_ = *g*) statistically unstable if not impossible.

This motivates us to take advantage of potential between-domain similarities. We achieve this aim by learning *G* + 1 conditional distributions pr(***X***_*i*_ | *Y*_*i*_ = *c, D*_*i*_ = *g*), *g* = 0, 1, …, *G*, in a data-driven way for causes *c* = 1, …, *C*, respectively. Consider any cause *c*, by Equation (2.4), pr(***X***_*i*_ | *Y*_*i*_ = *c, D*_*i*_ = *g*) is fully parameterized by (**Θ**^(*c*)^, ***λ***^(*c,g*)^). Because **Θ**^(*c*)^ is shared across domains, the *G* + 1 vectors of class-mixing weights {***λ***^(*c,g*)^, *g* = 0, 1, …, *G*} fully determines the between-domain differences in Equation (2.4). This points us to the strategy of encouraging *a priori* similar values of vectors ***λ***^(*c,g*)^ across domain *g* = 0, 1, 2, …, *G*; the degree to which they are similar may differ by cause. Data from all the domains can then be used to learn the degrees of optimal pooling between the domains for the *C* causes, respectively.

To achieve this aim, in Section 3.1.2, we introduce a tree-structured shrinkage prior for the *G* + 1 vectors of class-mixing weights {***λ***^(*c,g*)^, *g* = 0, 1, …, *G*} for each *c*. The prior is based on a logistic stick-breaking Gaussian process, diffused along a pre-specified rooted weighted domain tree with *G* + 1 leaves that encodes external between-domain similarity information. Also see Appendix E in the Supplementary Materials for a review of the general statistical strategy of specifying tree-structured shrinkage priors (Thomas *and others*, 2020; Li *and others*, 2021*a*).

#### 3.1.2 Logistic stick-breaking Gaussian diffusion prior for *λ*^(*c,g*)^: integrating domain hierarchy

Recall that 𝒯_*w*_ = (𝒯 = (𝒱, *E*), *w*) represents a rooted weighted tree over domains, where the *G* + 1 leaves 𝒟_leaf_ comprise *G* source domains and one target domain; see Section 5.2 for an example in the context of VA. Each domain comprises multiple independent observations {(***X***_*i*_, *Y*_*i*_) : *D*_*i*_ = *g*}. Each domain *g* is one-to-one mapped to a leaf in 𝒯_*w*_. We specify a prior based on a logistic stick-breaking Gaussian process diffused along 𝒯_*w*_ and end at *G* + 1 leaves, inducing a prior distribution over the class weights ***λ***^(*c,g*)^, *g* = 0, 1, …, *G*. We first reparameterize ***λ***^(*c,g*)^ with a stick-breaking representation: 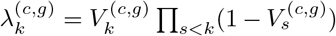, for *k* ∈ [*K*], where 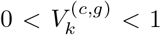, for *k* ∈ [*K* − 1] and 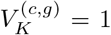. In particular, let 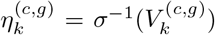, *k* ∈ [*K* − 1], *g* ∈ 𝒱_leaf_, where *σ*(*x*) = 1*/*{1+exp(−*x*)} is the sigmoid function. The logistic stick-breaking parameterization is completed by

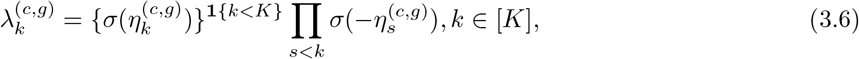

where **1**{*A*} is indicator function and equals 1 if statement *A* is true and 0 otherwise. This reparametrization lends itself to simple and accurate posterior inference via variational Bayes algorithms.

For a leaf *g* ∈ 𝒱_leaf_, let

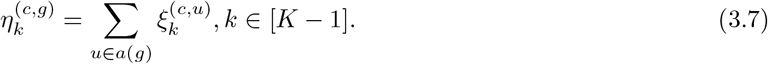

Note that 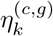 is defined for leaves *g* = {0} ∪ [*G*] only and 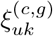 is defined for all the nodes *u* ∈ 𝒟. Finally, for *c* = 1, …, *C*, we specify

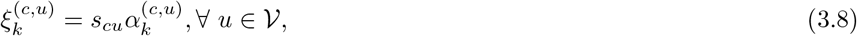

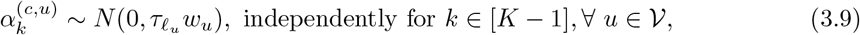

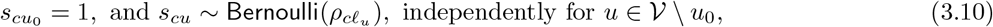

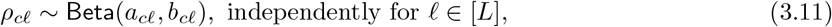

where *N* (*m ′, s ′*) represents a Gaussian with mean *m ′* and variance *s*^*′*^. In addition, *𝓁*_*u*_ ∈ [*L*] and maps node *u* ∈ 𝒟(leaf or non-leaf) to one of *L* levels; this enables distinct degrees of diffusion for *L* non-overlapping blocks of a pre-specified partition of the nodes. Let ***τ*** = (*τ*_1_, …, *τ*_*L*_)^T^ be the diffusion variances for *L* levels of nodes in the domain tree. Let ***s*** = {*s*_*cu*_, *c* ∈ [*C*], *u* ∈ 𝒟} be a *C* × *p* matrix of slab component indicators. Let ϱ be a *C* × *L* matrix with (*c, 𝓁*)-th element *ρ*_*c𝓁*_. Note that the spike-and-slab indicator probabilities 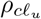 for a node *u* in the domain tree may vary by cause; relative to a more restrictive form 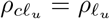, the present specification has the additional flexibility of cause-specific degree of pooling between domains. When *K >* 1 and *s*_*cu*_ = 1, ∀*u* ∈ 𝒱_leaf_, we would assume domains differ in class weights with probability one.

### 3.2 Prior for other parameters

We assume independent Dirichlet priors for the CSMFs in the source and target domains:

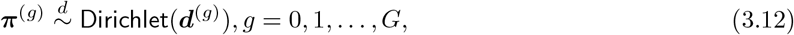

where 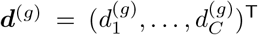 is a vector of hyperparameters; let ***d*** = {***d***^(*g*)^ : *g* = 0, 1, …, *G*}. In our simulations, we simply use ***d***^(*g*)^ = **1** to represent a uniform prior over all cause which works well empirically. In practice, informative knowledge can be incorporated by modifying these ***d***^(*g*)^ hyperparameters to match prior numbers of observed deaths of each cause in domain *g*.

#### 3.2.1 Secondary feature of the method: integrating cause hierarchy

In a particular analysis, the number of causes can be large. Causes considered to be similar in nature and etiology would produce a symptom with similar probabilities. In addition, the number of deaths due to a cause can be small, resulting in unstable estimation of the response profiles if done separately from other causes. To overcome these issues, {**Θ**^(1)^, …, **Θ**^(*C*)^} is also equipped with a tree-structured prior with a pre-specified cause tree of *C* leaves that encodes between-cause similarities. For example, Figure 3 shows a cause tree in our VA application (left margin) representing a hierarchy of 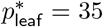 causes and 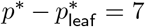 internal nodes that represent coarser aggregated causes; we specify all edge weights to be one. By doing so, we encourage *a priori* similar values of **Θ**^(*c*)^ across causes *c* = 1, …, *C*. This facilitates optimal pooling of information over causes and overcomes statistical stability issues for rare causes that would otherwise require ad hoc manual cause aggregations (Datta *and others*, 2021). Although the proposed approach can accommodate two hierarchies (domain and cause), because domain adaption is our primary goal, in the following we will focus empirical evaluations on the use of domain hierarchy.

Let 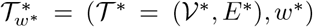 represent a rooted weighted tree, where the leaf set 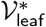 represents distinct causes of death labeled as *c* = 1, …, *C*. Each cause is mapped to one and only one leaf in 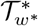. Note that ***Y*** maps every row of data **X** to a leaf in the tree for causes 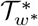 ; but this link is only observed for deaths occurred in the source domains {*i* : *D*_*i*_ *≠* 0}. Similarities between causes are then characterized by between-cause distances in 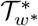. We then specify a logistic Gaussian diffusion prior:

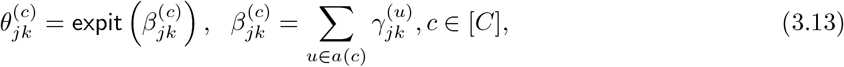

with Gaussian increments over the edges leading to each leaf:

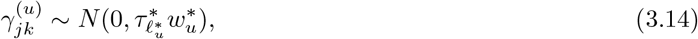

where 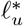 maps node *u* in the cause tree to one of *L*^*^ level; this is to allow distinct diffusion variances for *L*^*^ nonoverlapping blocks of a pre-specified partition of the nodes in the cause tree. Let 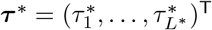 be the vector of diffusion variances for the *L*^*^ sets of nodes in the cause tree. Unlike in Section 3.1.2, we choose not to use node-specific spike-and-slab priors in the cause tree, which is equivalent to less aggressive shrinkage between causes and performs well in our simulation and validation studies.

Remark 3.1 Taken together, Sections 3.1.2 and 3.2.1 propose a prior for conditional distributions of the VA responses given any cause is a two-way tree-structured priors: i) the shrinkage among the domains in the columns is guided by a domain tree, and, ii) the shrinkage among the causes in the rows is guided by a cause tree. In particular, the shrinkage across causes is not domain-specific, but rather determined by information pooled across all domains. However, the shrinkage across domains is determined by a global-local structure, where we use 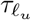 as diffusion variance parameter for all causes (“global”; 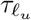 not indexed by *c*) and we use 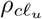 to introduce cause-specific (“local”; 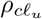 indexed by *c*) shrinkage of a leaf towards its parent (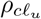 closer to 0 or 1 for stronger or weaker shrinkage).

### 3.3 Joint distribution

The joint distribution is fully specified by Equations (2.1-2.3), (3.6-3.11), (3.12), and (3.13-3.14). We collect the unobserved quantities by 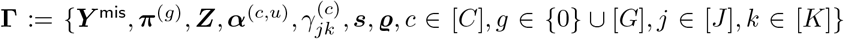.

We have the joint distribution of data 𝒟 and **Γ** as follows:

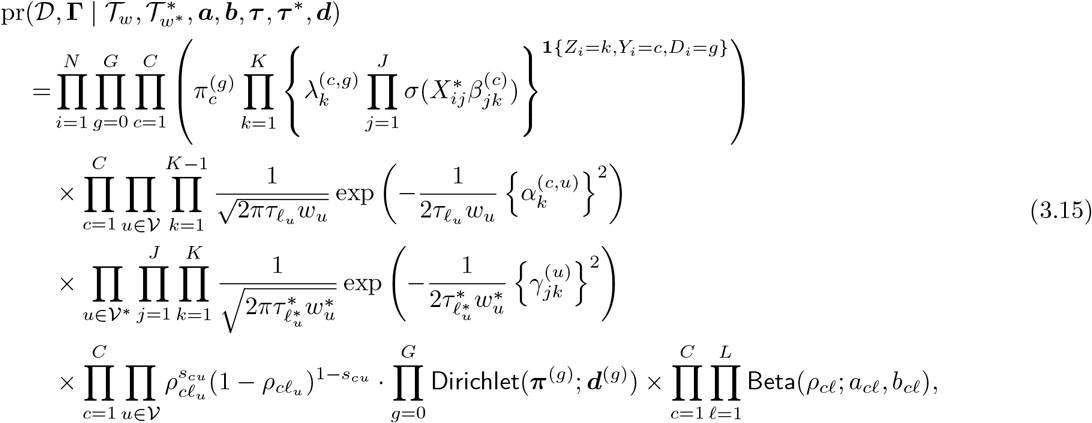

where Bernoulli likelihood components 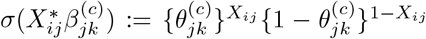 with 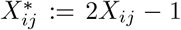. Our primary quantity of interest is the CSMFs in the target domain ***π***^(0)^ and the individual-specific cause-specific posterior probabilities 𝕡(*Y*_*i*_ = *c* | *D*_*i*_ = 0, 𝒟), *c* ∈ [*C*].

The directed acyclic graph (DAG) in Appendix Figure 2 in the Supplementary Materials shows the relationship between the observables, unknown quantities and hyper-parameters.

**Fig. 2:**
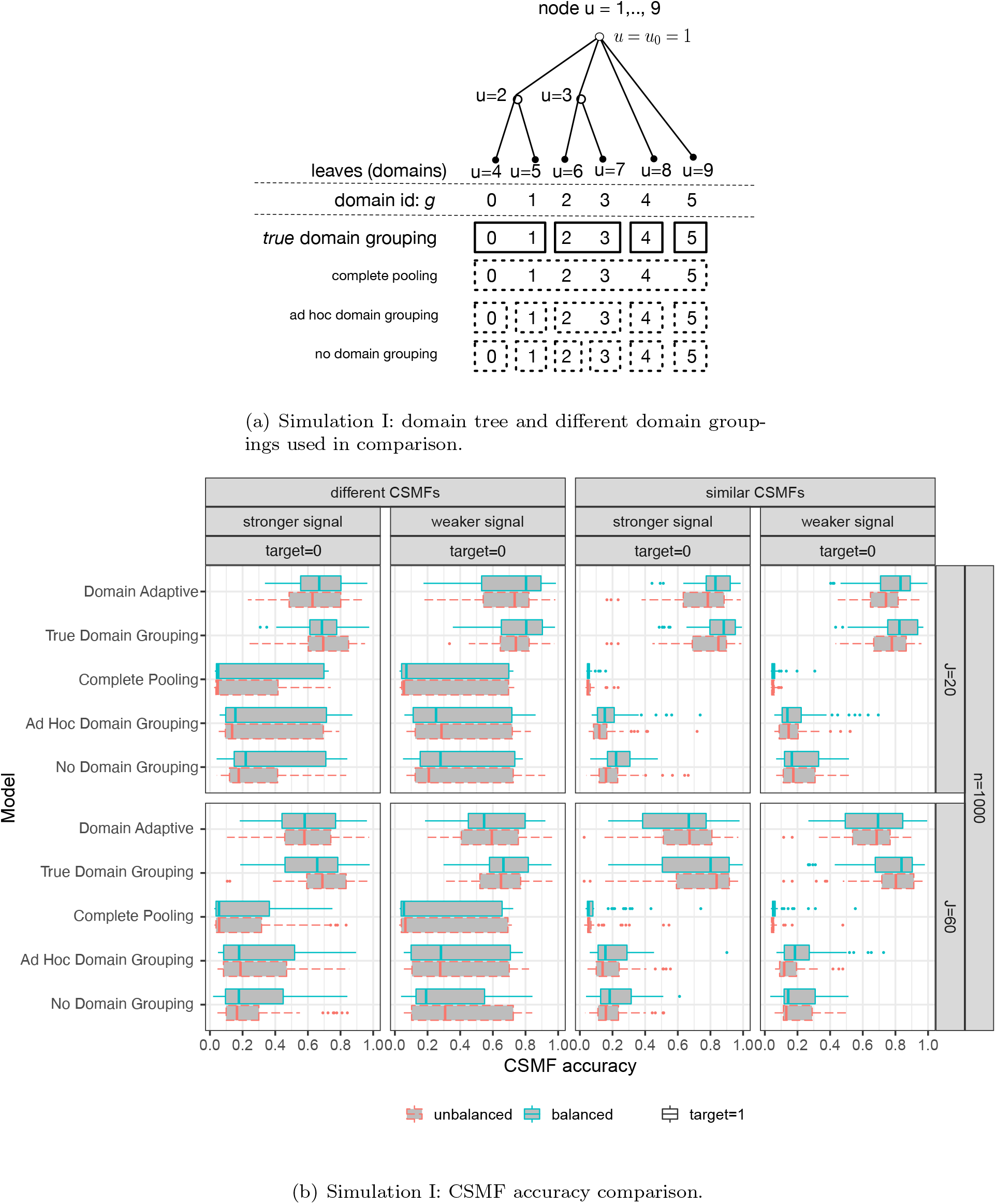
Simulation I: domain tree setup and results.

**Fig. 3:**
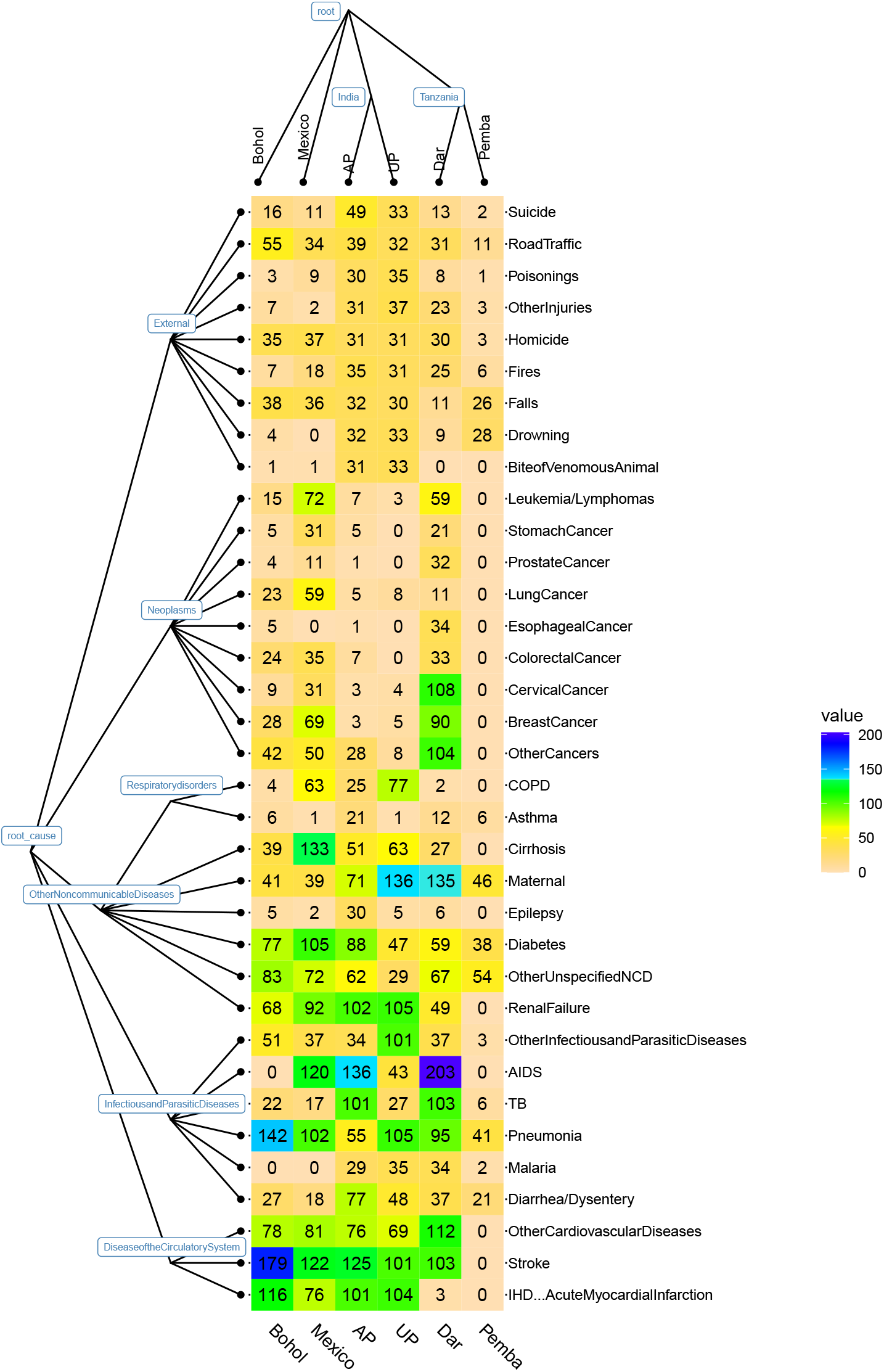
Death counts by cause and site for *N* = 7, 841 deaths and *J* = 168 across all six sites in the PHMRC data set. The exact death counts are shown in corresponding cells. Shown on the left and top margins are the cause and domain hierarchies assumed in the data analysis. We will mask the causes-of-death in one site during method testing so the site with masked causes is the target domain and the rest sites are source domains. The domains closer in the domain hierarchy (top margin) are *a priori* more likely fused to have the same vector of class mixing weights. The proposed method has a secondary feature that can incorporate a cause hierarchy (left margin) with 35 causes on the leaves and six aggregated causes represented by internal nodes.

## 4. Bayesian inference algorithms

Calculating a posterior distribution often involves intractable high-dimensional integration over the unknowns in the model. Traditional sequential sampling approaches such as Markov chain Monte Carlo (MCMC) remains a widely used inferential tool based on approximate samples from the posterior distribution. They can be powerful in evaluating multidimensional integrals. However, they do not guarantee closed-form posterior distributions. Variational inference (VI) is a popular alternative to MCMC for approximating the posterior distribution and has been widely used in machine learning and gaining interest in statistics (e.g., Blei, Kucukelbir and McAuliffe, 2017; Ormerod and Wand, 2010). In particular, VI has also been used for fitting the classical LCMs (e.g., Grimmer, 2011). VI requires a user-specified family of distributions that can be expressed in tractable forms while being flexible enough to approximate the true posterior; the approximating distributions and their parameters are referred to as “variational distributions” and “variational parameters”, respectively. VI algorithms find the best variational distribution that minimizes the Kullback-Leibler (KL) distance between the variational family and the true posterior distribution. VI has been widely applied in Gaussian (Carbonetto, Stephens *and others*, 2012; Titsias and Lázaro-Gredilla, 2011) and binary likelihoods (e.g., Jaakkola and Jordan, 2000; Thomas *and others*, 2020). Also see Blei, Kucukelbir and McAuliffe (2017) for a detailed review. We use VI because it is fast, bypasses infeasible analytic integration or data augmentation that is otherwise needed for MCMC under Dirac spike components and prior-likelihood non-conjugacy (Tüchler, 2008), and enables data-driven selection of hyperparameters via approximate empirical Bayes (Step 3, Appendix A in the Supplementary Materials).

We wish to obtain the marginal posterior distributions pr(***π***^(0)^ | 𝒟) and pr(*Z*_*i*_ | 𝒟). We conduct posterior inference via variational inference. We assume the variational distributions can factorize as follows:

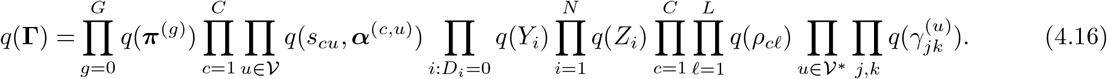

By the well-known equality, log pr(𝒟) = ε(*q*) + *KL*(*q*||pr(**Γ** | 𝒟)). Because log pr(𝒟) is constant in *q*, minimizing the KL divergence between the variational family and the true posterior distribution is equivalent to maximizing ε(*q*), or “evidence lower bound (ELBO)” which is defined by 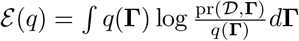 where **Γ** collects all the unknowns. We further bound ε(*q*) from below by bounding terms in pr(𝒟, **Γ**) that involve sigmoid functions that create non-conjugacy issues under the Gaussian-distributed priors used in this paper hindering simple closed-form VI updates. In particular, following Jaakkola and Jordan (2000), we can bound

Equation (3.6) and 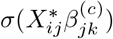 from below respectively by

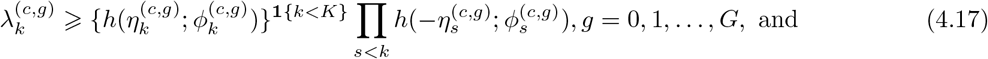

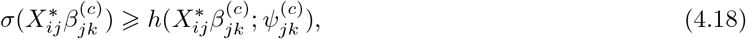

where we have used the inequality

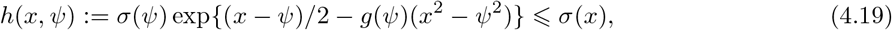

with 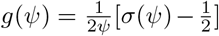 where *ψ* is a tuning parameter. As a result, the right-hand-side terms in Equations (4.17) and (4.18) are quadratic in 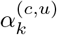 and 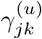, paving the way for closed-form VI updates. Also see Durante, Rigon *and others* (2019) for a modern view of the technique as a bona fide variational algorithm with Pólya-Gamma augmentation. We now have a lower bound ε*(*q*) of ε(*q*) which is defined as

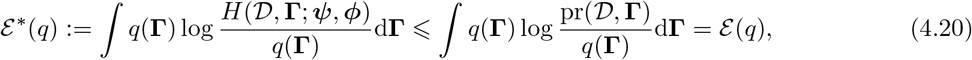

where *H ⩽* pr(𝒟, **Γ**) is obtained by applying the lower bounds in Equations (4.17) and (4.18) to relevant terms in (3.15) and has tuning parameters 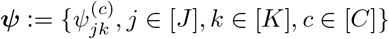 and 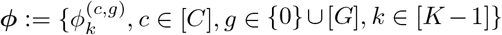; see Appendix B in the Supplementary Materials for the exact formula for calculation.

The variational algorithm then finds the optimal variational distribution in the variational family that maximizes ε*(*q*). In particular, we take the logarithm of the lower bound *H* of the joint probability density for data and unknowns with respect to a variational distribution *q*: 𝔼_*q*_[log *H*]. The algorithm updates each factor in order while holding the rest fixed. The update for the *j*-th factor in the variational distribution is 𝔼_*q−j*_ [log *H*], where 𝔼_*q−j*_ means taking expectations with respect to *q* over all but the variables in the *j*-th factor in *q*. The logarithmic of *H* can be written as in Appendix B in the Supplementary Materials. A desirable property of log *H* is that integration of log *H* with respect to each factor of *q* is in closed-form, which is a key ingredient of each VI update. The pseudo-code for the VI updates are provided in Algorithm 1. See Appendix A in the Supplementary Materials for the details of each update.

### Cause-specific domain dissimilarity measure

The framework motivates the following the estimated cophenetic distance (e.g., Sneath, Sokal *and others*, 1973) between the target domain and each of the source domains. In particular, the dissimilarity measure is 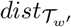 (`target` = 0, `source` = *g*; `cause` = *c*), *g* = 1, …, *G, c* = 1, …, *C*, where *w* (*P*) is defined as the sum of all the modified edge weights along the path *P* connecting the target domain and source domain *g* in 𝒟_*w′*_, and the modified weight of an edge (*pa*(*u*) → *u*) is 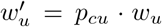 for node *u* ∈ 𝒟in the domain tree (see Equation (A5) of VI updates in Appendix A in the Supplementary Materials for definition of *p*_*cu*_ (on a logit scale) as the variational approximation to ℙ (*s*_*cu*_ = 1 | 𝒟)).

*Choice of K* We follow Bishop (2006) and use criterion 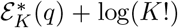 where 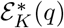 is the lower bound of log marginal data likelihood for a *K*-class model and the correction term is to make different models comparable (e.g., Grimmer, 2011, Section 5.2).

*Software* A free and publicly available R package that implements the VI algorithm for scalable approximate posterior inference is freely available at https://github.com/zhenkewu/doubletree. The package is designed to work under all possible patterns of observed and missing causes of death: (`Scenario i`) *Y*_*i*_ is missing in a single domain (say *g*): i-1) none has confirmed causes in domain *g*; i-2) there exists at least one observed *Y*_*i*_ in domain *g*; (`Scenario ii`) *Y*_*i*_ is missing in *M* ⩾ 2 domains; (Scenario iii) no missing *Y*_*i*_ in any domain. This paper has focused on Scenario (i-1) for simpler exposition.

## 5. Simulation studies

We conduct two sets of simulation studies to evaluate the operating characteristics of the proposed method and demonstrate its better capability of estimating the target-domain CSMFs and assigning individual-level causes of death relative to a few alternatives with ad hoc specifications of information pooling across the domains. In the first set of simulations, we simulate data based on true parameters values under NLCM. In the second set of simulations, we use a validation data set and selectively mask a subset of deaths’ true causes in a synthetically constructed target domain and then apply NLCM. The simulation designs, performance metrics, and results are detailed below.

### Performance Metrics

First, we assess the overall accuracy by the so-called “CSMF accuracy” (Murray *and others*, 2011*b*), widely used as the metric to compare the estimated CSMF vector against the truth. The CSMF accuracy metric measures the *L*_1_-distance between the estimated and true vectors of CSMFs, and is normalized to range between 0 (worst) and 1 (best). It is defined as 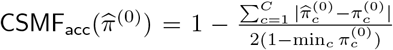, where 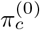 is the true CSMF for cause *c* that we set in simulation design (or calculated by the empirical distribution in the synthetic target domain data in Simulation II below). This formulation is also known as the normalized absolute error in the quantification learning literature (González *and others*, 2017). Finally, for the accuracy of COD classifications, we will use top cause accuracy: the fraction of deaths with the true CODs in the top predicted causes.

### 5.1 Simulation I

#### Design

We simulated *R* = 200 independent replicate data sets for different total sample sizes (*N* = 1000, 4000). To illustrate, we use a domain tree 𝒯_*w*_ shown in Figure 2(a) with equal edge weights and true domain leaf groups; 𝒯_*w*_ has *p*_leaf_ = 6 leaves and 3 domain leaf groups. The leaf “0” is set to be the target domain leaf. For each *N*, we set each domain’s sample size to be approximately *N/p*_leaf_ for *g* = 0, 1, …, *G* (with rounding where needed) to investigate balanced leaves and set the sample size in domain *g* to be approximately 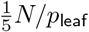 or 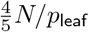 with equal chances for mimicking unbalanced observations across the domain leaves. Within domain *g*, we further assign deaths into *C* causes by independently sampling from categorical distributions with CSMFs ***π***_*g*_, *g* = 0, 1, …, *G*. We then simulated multivariate binary response data for different dimensions *J* = 20, 60, for *K* = 2 classes according to Equations (2.2) and (2.3). We considered *C* = 3 causes. Two different sets of {**Θ**^(*c*)^} were considered; see Appendix D in the Supplementary Materials for more details of the true parameter values and model setup.

For each simulated data set, we fitted the proposed model, based on which we compute the approximate posterior mean of ***π***^(0)^ obtained via its optimal variational distribution. In addition, we also compared against a few NLCM-based approaches but with suboptimal, ad hoc specifications of information pooling between the domains (to different degrees of cross-domain shrinkage). Figure 2(a) shows the domain groupings used in these comparisons. In summary, we fit 1) the proposed method:”`Domain Adaptive`”; 2) “True Domain Grouping”: for any cause *c*, assume identical pr(***X***_*i*_ | *Y*_*i*_ = *c, D*_*i*_ = *g*) for domains in a group (4 groups in the simulation truth). To do so, for each *c*, we fix *s*_*cu*_ to 1s or 0s in a way that results in the true domain grouping; 3) “`Complete Pooling`”: completely ignore the external domain tree information during estimation and also ignore the sample-to-domain mappings ***D***. By doing so, we assume pr(***X***_*i*_ | *Y*_*i*_ = *c, D*_*i*_ = *g*) remains the same between the domains; 4) “`Ad hoc Domain Grouping`”: use a manual domain grouping that is finer than the true domain grouping; 5) “`No Domain Grouping`”: same as 3) except ***D*** is used during estimation so that data are recognized to have come from different domains.

#### Results

Figure 2(b) compares the methods in terms of CSMF accuracy. The proposed NLCM Domain Adaptive method adaptively learns the the domain groupings and produced the CSMF estimate for the target domain with the highest accuracy. The accuracy is comparable to the ones obtained under the true domain grouping. The method with complete pooling between domains generally performs the worst; this should not be surprising given the simulation truth is to mimic situations where pr(*X*_*i*_ | *Y*_*i*_, *D*_*i*_ = *g*) differs by domain. NLCMs with ad hoc domain grouping and no grouping produced more accurate estimates than the one with complete pooling between the domains. However, because both methods are based on domain groupings that are finer than the true domain grouping, they do not fully use similar domains to improve the accuracy of estimating the conditional distributions of the VA responses given a cause, resulting in sizable losses in CSMF accuracy. Similar relative patterns are also clear when RMSEs are compared (see Figure Appendix Figure 3). In addition, as expected the conditional dependence modeled by the NLCMs for each cause improved the individual-level classification performance (top cause accuracy) relative to methods that ignored conditional dependence (results not shown here).

### 5.2 PHMRC VA Data: Background and Description

In Section Appendix D.1 and Section 6, we will use the Population Health Metrics Research Consortium (PHMRC) VA validation data with physician-coded causes of death to evaluate the performance of the proposed method. We first review aspects of the data set pertinent to our study.

PHMRC VA validation data collection was implemented in six sites in four countries: Andhra Pradesh (“AP”), India; Uttar Pradesh (“UP”), India; Dar es Salaam (“Dar”), Tanzania; Pemba Island (“Pemba”), Tanzania; Bohol, Philippines; Mexico City (“Mexico”), Mexico. The goal is to create a high quality validation data set from different populations to evaluate comparative method performance and make recommendations for future VA implementations. See Murray *and others* (2011*a*) for a more complete description of the PHMRC VA validation data. The data set with 7,841 adult deaths and 168 symptoms that we use here is further based on preprocessing in McCormick *and others* (2016).

Figure 3 shows the domain hierarchy via a rooted tree with six leaves at the top margin. The domain hierarchy uses country membership information to form the leaves and internal nodes. Unless otherwise stated, we set all edge weights to be one for illustration. External domain-level information that is highly associated with CSMFs may also be incorporated to form the domain hierarchy, e.g., via hierarchical clustering of a variety of domain-level information that may alter symptom-cause relationships, such as time periods, level of VA interviewer training, and differential availability of treatments that mitigate a subset of symptoms interviewed in VA. As a secondary feature of the proposed method, the left margin of Figure 3 shows the cause hierarchy with 35 leaves along with coarser aggregated cause definitions.

Among the 168 symptoms, 63 have a missing rate of higher than 1%, of which 37 has a missing rate higher than 5%. The highest missing rate is 96.6% for: “Was there pain in the upper belly?”; the next highest missing rate is 92%, for four questions related to where rash was located if present: “Trunk?”,”Extremities?”,”Everywhere?”, “Other locations?” In addition, the numbers of missing symptoms for a subject are between 1 and 76, with a median of 22. Missing data in VA in the form of “Don’t Know” or “Choose not to answer” appear for various reasons. Missing data have been considered by Kunihama *and others* (2020) under the assumption of missing at random. Absent additional information regarding individual-specific missing data mechanism and given the lack of likely alternative sensitivity assumptions about missing data, we will assume missing at random in this paper when conducting model estimation and method comparisons. This sets the stage for fair relative comparisons with other existing methods that either assume missing completely at random or missing at random. In our proposed model, because given a cause and a class, symptoms are assumed mutually independent, when calculating the cause-specific likelihood for an individual in class *k* with a subset of missing symptoms, we simply do products of Bernoulli likelihoods only over symptoms with non-missing information; see the implementation in Steps 1a and 1e in Appendix A during variational updates.

### 5.3 Simulation II: Semi-Synthetic

Here we use the PHMRC VA validation data to evaluate the performance of the proposed method. Because validation data contains gold-standard causes of death, we split the original data set into source and target domain data sets and mask the causes of the deaths allocated to the target domain. Because we use the real PHMRC data while masking a subset of causes of death, we refer to this simulation as “semi-synthetic”. See Appendix D.1 in the Supplementary Materials for the details about the design and results showing that domain adaptive estimation provides more accurate CSMF estimates and top cause COD assignment.

## 6. Domain Adaptation across Actual PHMRC Sites

This scenario uses the actual PHMRC domain designation: one site as the target domain with the gold-standard cause-of-death labels masked, the rest five sites as source domains with observed gold-standard causes-of-death. Table 1 shows the CMSF accuracy when each of the six PHMRC study sites is treated as the target domain iteratively. The method selected two-class models. The domain adaptive approach achieved better accuracies in CSMF estimation and slight improvements on the top-cause classification accuracies. The somewhat low accuracies using PHMRC data are well known, motivating new ongoing validation data collection by our substantive collaborators based on which we will further test our method. To illustrate how to interpret results from the domain adaptive method, we pick AP as the target domain. In addition, we pick six causes (out of 35 causes used during model fittings) to illustrate the results of cause-specific shrinkage. The causes are selected based on the different levels of shrinkage: near-complete pooling (cause “Drowning”), and no substantial shrinkage between the domains (cause AIDS, Stroke, Renal Failure, Tuberculosis (TB), Inflammatory Heart Disease (IHD)-Acute Myocardial Infarction (MI)).

**Table 1:** CSMF accuracy when each of the six PHMRC study sites is treated as the target domain iteratively.

| Method | Bohol | Mexico | AP | UP | Dar | Pemba |
| --- | --- | --- | --- | --- | --- | --- |
| *CSMF Accuracy* |  |  |  |  |  |  |
| Domain Adaptive | 0.68 | 0.72 | 0.70 | 0.66 | 0.66 | 0.63 |
| Complete Pooling | 0.67 | 0.67 | 0.67 | 0.61 | 0.61 | 0.58 |
| Ad Hoc Domain Grouping | 0.67 | 0.68 | 0.68 | 0.61 | 0.65 | 0.62 |
| No Domain Grouping | 0.67 | 0.64 | 0.66 | 0.62 | 0.63 | 0.60 |
| *Top Cause Accuracy* |  |  |  |  |  |  |
| Domain Adaptive | 0.32 | 0.30 | 0.34 | 0.36 | 0.34 | 0.41 |
| Complete Pooling | 0.32 | 0.29 | 0.35 | 0.36 | 0.33 | 0.39 |
| Ad Hoc Domain Grouping | 0.32 | 0.30 | 0.36 | 0.35 | 0.33 | 0.38 |
| No Domain Grouping | 0.32 | 0.30 | 0.34 | 0.35 | 0.33 | 0.40 |

Using AP as an example target domain, Figure 4 shows two-class LCM results for some pairs of (cause,domain). For each of the six causes shown in Figure 4(b), the relative importance of the two classes varies greatly by domain, indicating substantive differences in pr(***X***_*i*_ | *Y*_*i*_ = *c, D*_*i*_ = *g*) between the domains. This is evident from the patterns of the class mixing weights for “TB” and “Renal Failure”. Interestingly, this is not uniformly true for all causes, mostly notably for “Drowning”, which placed large weights on the first class regardless of domain. A plausible explanation for this empirical finding is that symptoms are easily recognizable and highly specific if a death is caused by drowning. This analysis also empirically confirms that the conditional distributions of VA responses given each cause may vary by cause.

**Fig. 4:**
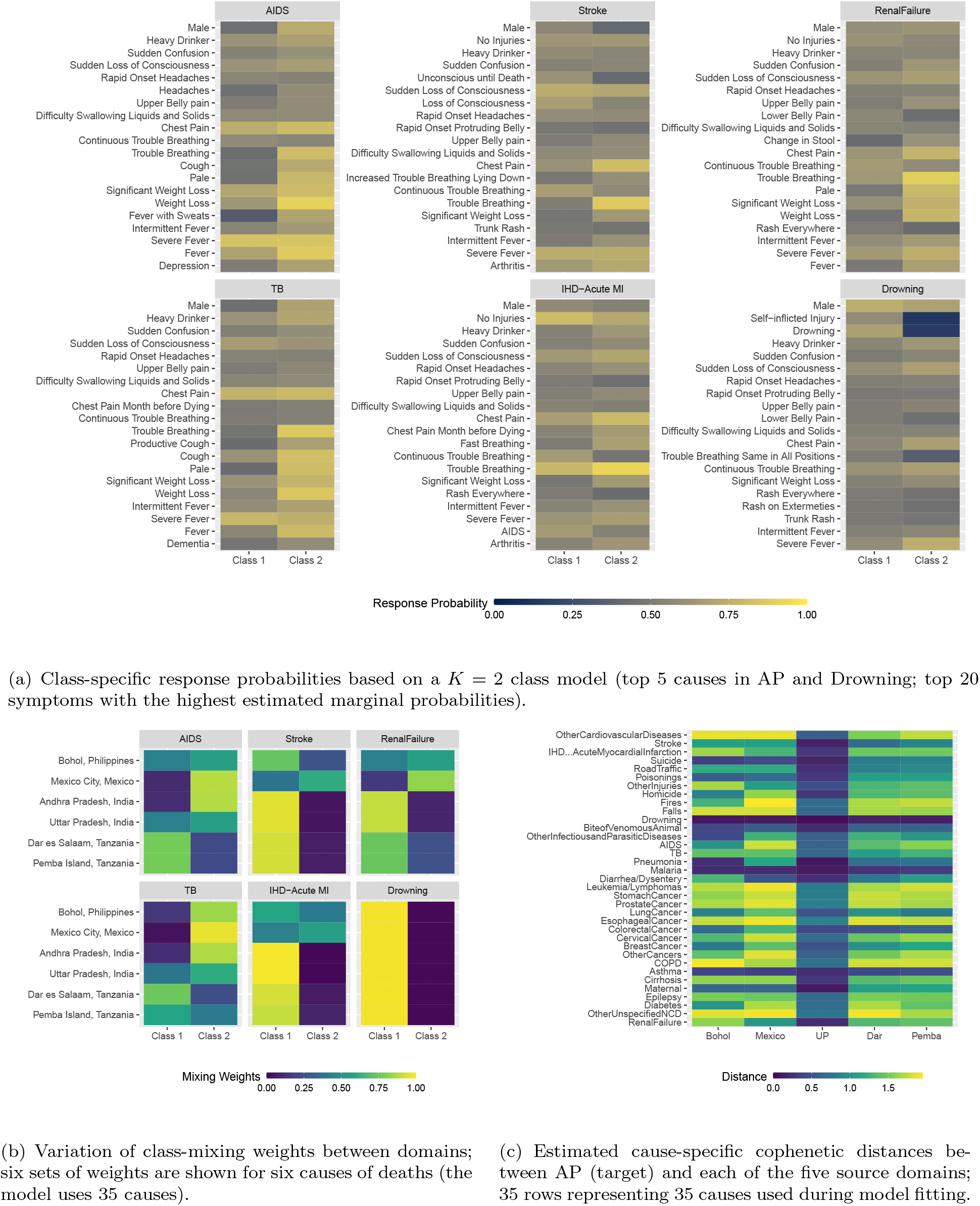
PHMRC results based on a *K* = 2 class nested LCM; for illustration, model results using AP as the target domain are shown here.

In addition, we also calculate the estimated cophenetic distance (e.g., Sneath, Sokal *and others*, 1973) between the target domain and each of the source domains (see Section 4). Figure 4(c) shows for each of 35 causes (rows), five estimated distances with smaller values representing higher similarity between the target domain (AP) and each of the five source domains shown on the x-axis. We make a few interesting empirical observations. First, UP is estimated to be most similar to the target domain for almost all causes, which is perhaps unsurprising given AP and UP are two states in India. Second, the joint pattern of similarity between AP and five source domains differ by cause. For example, for causes like “Drowning”, “Malaria”, “Asthma”, the dissimilar measures are all estimated to be small, indicating AP is uniformly similar to all other source domains in terms of symptom-cause conditional distributions. This uniformity in the dissimilarity measure may be explained by reporting of specific symptoms that vary little by domain. On the other hand, for causes with complex etiologies and manifest symptoms such as “Renal Failure”, the target domain AP is estimated to be most similar to UP and least so to Bohol.

## 7. Discussion

### Summary

In this paper, we presented a hierarchical Bayesian approach to use individual-level multivariate binary responses obtained from a target domain in the absence of any gold-standard categorical labels (“causes” in VA) for the estimation of the target population fractions of the label categories (“CSMFs” in VA). This is made possible by using individual-level data from multiple source domains where additional gold-standard cause labels are available. The data from multiple domains are integrated following a data-driven tree-structured shrinkage approach, so that for each cause, domains that have similar conditional distributions of the responses given the cause are encouraged to be pooled to improve estimation. We achieved this goal via a logistic stick-breaking Gaussian diffusion process on the mixing weights along a pre-specified domain hierarchy. In addition, an analyst may use another hierarchy applied to causes to regularize the parameter estimation when characterizing the conditional distributions of the responses given a cause. Simulation studies show that the proposed method produces more accurate estimation of CSMFs than methods that either ignore between-domain differences by complete pooling of data across the domains, ad hoc specification of domain grouping, or no domain grouping at all; the proposed method performs similarly to an oracle method with the true domain groupings. Although this paper focuses on the more challenging case where no cause label is observed in the target domain for simpler exposition, the model readily generalizes to using gold-standard cause labels for a subset of deaths in the target domain. The software accompanying this paper has implemented such extensions.

### Limitations

Practical limitations of the proposed approach may exist. First, the hierarchy for the domain is pre-specified based on external domain-level information (geo-locations of the study sites in this paper) and are not estimated from the VA data themselves. Methods that specify a prior over the space of domain hierarchies for posterior inference may be fruitful at extra computational costs (e.g., Knowles and Ghahramani, 2014). Second, deviation from missing-at-random assumption for the VA questionnaire responses may impact the model results and performance during domain adaptations. This issue remains challenging and to be explored in collaboration with VA substantive experts to identify common and major reasons leading to missing data. Third, the cause list used in this paper is chosen based on clear clinical meanings of distinct etiologic implications and also for methodological illustration. Certain causes of death, such as COVID-19 related deaths, may quickly emerge as prominent causes in some populations that necessitates updated cause lists. Fourth, PHMRC data is currently the only validation data set for evaluation; future VA gold standard data sets will be available for additional validation. Fifth, additional unstructured narrative texts are available, and may improve the capacity of the present approach by augmenting the VA symptoms with absence or presence of derived text features (e.g., cause-discriminative words). Finally, the current results are agnostic to additional information about symptom-cause relationships. Estimation accuracy may be further improved by incorporating these information (e.g., McCormick *and others*, 2016; Schifeling, Reiter *and others*, 2016).

### Future directions

There are a few statistical extensions that may further improve the utility of the proposed method. First, additional individual-level covariates, such as age, pregnancy status, and seasonality, that may explain variation in the conditional distribution of the VA questionnaire responses given a cause pr(***X***_*i*_ | *Y*_*i*_, *D*_*i*_). Our framework readily incorporates discrete individual-level covariates via concatenation with the vector of VA questionnaire responses. For continuous individual-level covariates, Moran *and others* (2021) illustrated an approach based on a different framework of factor models, which however is not suited to dealing with a domain absent any cause-of-death label. Mixed outcome extensions are desirable (e.g., Zhang *and others*, 2021). Second, extensions that incorporate priors over *K* that may differ by cause can lead to posterior inference of cause-specific values of *K*. To this end, for each cause, sparsity priors over a probability simplex may be introduced to encourage absence of a subset of classes in certain domains. Third, latent class model for pr(***X***_*i*_ | *Y*_*i*_, *D*_*i*_) is a simple example of probabilistic tensor decomposition for multivariate discrete data, which can be replaced with alternatives against which comparisons are warranted (e.g., Bhattacharya and Dunson, 2012; Zhou *and others*, 2015; Gu *and others*, 2021). Fourth, negative transfer issues have been noted in machine learning literature on transfer learning (e.g., Pan and Yang, 2009; Zhang *and others*, 2020). Our approach is based on an assumption of a shared set of class-specific response profiles **Θ**^(*c*)^ for each cause to facilitate interpretation. It is of interest to evaluate potential impact of deviations from such an assumption and to study mitigating solutions (e.g., Stephenson, Herring and Olshan, 2020). We leave these topics for future research.

## Supporting information

Details of the algorithm, additional simulation results

## Data Availability

All data produced are available online at https://github.com/zhenkewu/doubletree

http://ghdx.healthdata.org/record/ihme-data/population-health-metrics-research-consortium-gold-standard-verbal-autopsy-data-2005-2011

## Supplementary Material

Supplementary material is available online.

## Acknowledgments

This work is partly supported by a seed grant from Michigan Institute of Data Science (MIDAS; to ZW, IC, ML). *Conflict of Interest* : None.

## Algorithm 1

Pseudocode of Variational Bayes Algorithm

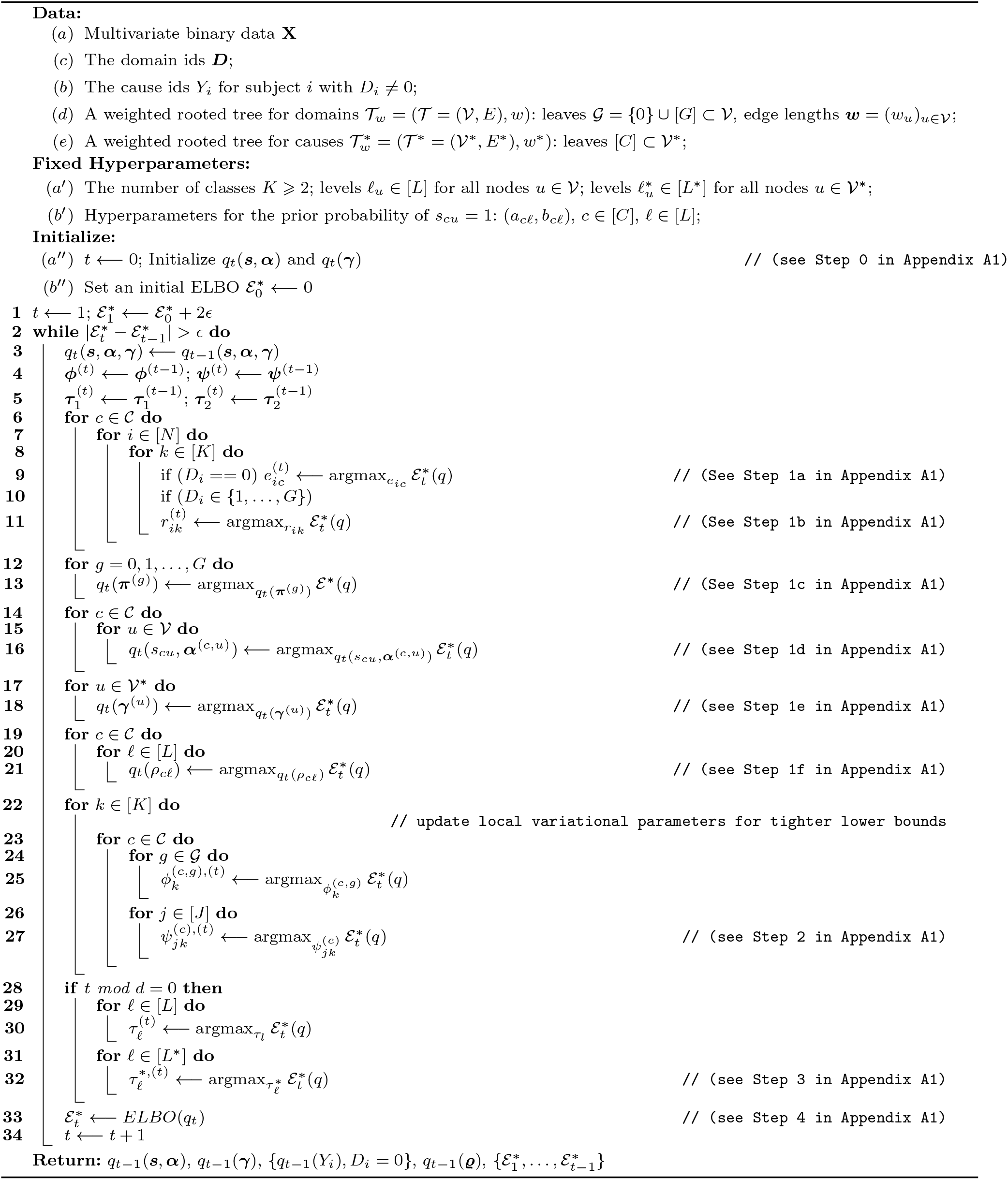

