## Supplementary material for "Tree-informed Bayesian multi-source domain adaptation: cross-population probabilistic cause-of-death assignment using verbal autopsy": Details of the algorithm, additional simulation results

#### Appendix A. Details of the Variational Inference Algorithm

In the following, let $q_t(A)$ represent a generic variational distribution for unknown quantities in $A$ at iteration $t$; Let $q_t(-A)$ represent the variational distribution for all but the random quantities in $A$. Let $\mathrm{pr}(A)$ represent a generic true joint distribution of the quantities in $A$. $[Q] := \{1, \ldots, Q\}$ represents the set of positive integers smaller than or equal to a positive integer $Q$. The algorithm presented below deals with missing data (under missing-at-random assumption for elements of $\boldsymbol{X}_i$ given the causes). Let $\mathcal{J}_i \subseteq \{1, \ldots, J\}$ denote the index set for the subset of observed responses for subject $i$. Let $\mathcal{I}_j \subseteq \{1, \ldots, J\}$ be the index set for the subset of subjects with observed $j$-th response. Finally, recall transformed response is $X^*_{ij} = 2X_{ij} - 1$; $\sigma(\bullet)$ denotes sigmoid function: $\sigma(x) = 1/(1 + \exp(-x))$.

*Step 0.* Initialize the variational distribution $q_t(\cdot)$ at $t = 0$. The update of each component of the variational distribution in Equation (4.16) of the Main Paper has a closed form that is determined by relevant first and second moments. We initialize these moments to initialize $q_0(\cdot)$. In addition, because the sigmoid functions are bounded by Gaussian kernels that depend on additional tuning parameters $(\boldsymbol{\psi}, \boldsymbol{\phi})$, we need to initialize them too. Finally, we initialize hyperparameters $(\boldsymbol{\tau}, \boldsymbol{\tau}^*)$.

In particular,

- Additive components of the logistic stick-breaking parameters $\alpha_k^{(c,u)}$ given $s_{cu} = 1$:

*To whom correspondence should be addressed.

$\{(\mu_{\alpha_k^{(c,u)},1}, \sigma^2_{\alpha_k^{(c,u)},1}) := (E_{q_t}\{\alpha_k^{(c,u)} \mid s_{cu}=1\}, V_{q_t}\{\alpha_k^{(c,u)} \mid s_{cu}=1\}) : k \in [K-1], c \in [C], u \in \mathcal{V}\}$. The mean and variance fully determine the optimal variational distribution for $\alpha_k^{(c,u)}$ given $s_{cu} = 1$, which can be shown to be a Gaussian distribution;

- Logit-transformed response probabilities: $\{(\mu_{\gamma_{jk}^{(u)},1}, \sigma^2_{\gamma_{jk}^{(u)},1}) := (E_{q_t}\{\gamma_{jk}^{(u)}\}, V_{q_t}\{\gamma_{jk}^{(u)}\}) : j \in [J], k \in [K], u \in [C]\}$;
- Tuning parameters in the Jaakkola-Jordan lower bounding technique: $\{\psi_{jk}^{(c)},\ j \in [J], k \in [K]\}$, $\{\phi_k^{(c,g)},\ c \in [C],\ g \in \{0\} \cup [G],\ k \in [K-1]\}$, and
- The hyperparameters $\{\tau_\ell, \ell \in [L]\}$, $\{\tau_\ell^*, \ell \in [L^*]\}$.

Compute additional first and second moments as follows:

$$
\begin{aligned}
E_{q_t}\left\{\eta_k^{(c,g)}\right\}^2 &= \sum_{u \in a(g)} \left\{ p_{cu}\left(\sigma^2_{\alpha_{k,1}^{(c,u)}} + (1-p_{cu})\left\{\mu_{\alpha_{k,1}^{(c,u)}}\right\}^2\right)\right\} + E^2_{q_t}[\eta_k^{(c,g)}],\\
E_{q_t}\left\{\alpha_k^{(c,u)}\right\}^2 &= p_{cu}\left(\sigma^2_{\alpha_{k,1}^{(c,u)}} + \left\{\mu_{\alpha_{k,1}^{(c,u)}}\right\}^2\right) + (1-p_{cu})\sigma^2_{\alpha_{k,0}^{(c,u)}},
\end{aligned}
$$

where $\sigma^2_{\alpha_{k,0}^{(c,u)}} = \tau_{\ell_u} w_u$ is the variance of $\alpha_k^{(c,u)}$ in its variational distribution given $s_{cu} = 0$ (as will be readily seen in Step 1d below according to the VI update for $\alpha_k^{(c,u)}$). Similarly, for the quantities in the cause hierarchy, we compute

$$
\begin{aligned}
E_{q_t}\left\{\beta_{jk}^{(c)}\right\}^2 &= \sum_{u \in a(c)} \sigma^2_{\gamma_{jk,1}^{(u)}} + E^2_{q_t}\{\beta_{jk}^{(c)}\},\\
E_{q_t}\left\{\gamma_{jk}^{(u)}\right\}^2 &= \sigma^2_{\gamma_{jk,1}^{(u)}} + \left\{\mu_{\gamma_{jk,1}^{(u)}}\right\}^2.
\end{aligned}
$$

Finally, compute $E_{q_t}\{\eta_k^{(c,g)}\} = \sum_{u \in a(g)} E_{q_t}\{\xi_k^{(c,u)}\}$, $E_{q_t}\{\xi_k^{(c,u)}\} = E_{q_t}\{s_{cu}\alpha_k^{(c,u)}\} = \mu_{\alpha_{k,1}^{(c,u)}}$.

$E_{q_t}[\beta_{jk}^{(c)}] = \sum_{u \in a(c)} E_{q_t}[\gamma_{jk}^{(u)}] = \sum_{u \in a(c)} \mu_{\gamma_{jk,1}^{(u)}}$.

Set initial $\mathcal{E}^*(q) = 0$.

At Step $t+1$, iterate between Step 1 to 4 until convergence (we omit iteration step index "$t$" and "$t+1$" in the notations below for simplicity):

*Step 1a.* Update $q_{t+1}(Y_i)$ for $\{i : D_i = 0\}$, by a categorical distribution with probabilities $\boldsymbol{e}_i = (e_{i1}, \ldots, e_{iC})^\mathsf{T}$:

$$
e_{ic} \propto \exp\left(E_{q_t}[\log \pi_c^{(0)}] + \sum_{k=1}^{K} r_{ik} F_{ik}^{(c,0)}(q_t)\right),
$$

where

$$F_{ik}^{(c,g)}(q_t) = \sum_{m<k} \left( \log\sigma(\phi_m^{(c,g)}) + \left\{-E_{q_t}(\eta_m^{(c,g)}) - \phi_m^{(c,g)}\right\}/2 - g(\phi_m^{(c,g)}) \left\{ E_{q_t}\left\{\eta_m^{(c,g)}\right\}^2 - \left\{\phi_m^{(c,g)}\right\}^2 \right\} \right)$$
$$+ \mathbf{1}\{k<K\} \left( \log\sigma(\phi_k^{(c,g)}) + \left\{E_{q_t}(\eta_k^{(c,g)}) - \phi_k^{(c,g)}\right\}/2 - g(\phi_k^{(c,g)}) \left\{ E_{q_t}\left\{\eta_k^{(c,g)}\right\}^2 - \left\{\phi_k^{(c,g)}\right\}^2 \right\} \right)$$
$$+ \sum_{j\in\mathcal{J}_i} \log\sigma(\psi_{jk}^{(c)}) + (X_{ij}^* E_{q_t}(\beta_{jk}^{(c)}) - \psi_{jk}^{(c)})/2 - g(\psi_{jk}^{(c)}) \left\{ E_{q_t}\left\{\beta_{jk}^{(c)}\right\}^2 - \left\{\psi_{jk}^{(c)}\right\}^2 \right\}, \tag{A1}$$

for $c \in [C]$ and $g \in \{0\} \cup [G]$. In addition, for observations with observed $Y_i = c$ we set $e_{ic} = 1$ and $e_{ic'} = 0$ for $c' \neq c$.

*Step 1b.* Update $q_{t+1}(Z_i)$ by a categorical distribution with probabilities $\boldsymbol{r}_i = (r_{i1}, \ldots, r_{iK})^\mathsf{T}$:

$$r_{ik} \propto \exp\left( \sum_{c=1}^{C} e_{ic} \left\{ F_{ik}^{(c,g)}(q_t) \right\} \right).$$

*Step 1c.* Update $q_{t+1}(\boldsymbol{\pi}^{(g)}), g \in \{0\} \cup [G]$ by

$$q_{t+1}(\boldsymbol{\pi}^{(g)}) \propto \mathsf{Dirichlet}\left( \sum_{i=1}^{N} e_{i1} + d_1^{(g)}, \ldots, \sum_{i=1}^{N} e_{iC} + d_C^{(g)} \right). \tag{A2}$$

*Step 1d.* Update $q_{t+1}(s_{cu}, \boldsymbol{\alpha}^{(c,u)})$ for each node $u \in \mathcal{V}$ of the tree $\mathcal{T}$ over the $G+1$ domains, which takes a form of two-component Gaussian mixture, separately for each cause $c \in [C]$. In particular,

$$\log q_{t+1}(s_{cu}, \boldsymbol{\alpha}^{(c,u)}) = \mathbb{E}_{q_t(-(s_{cu}, \boldsymbol{\alpha}^{(c,u)}))} \log H + \text{const}$$
$$= s_{cu} \sum_{k=1}^{K-1} \log\mathcal{N}(\alpha_k^{(c,u)}; \mu_{\alpha_k^{(c,u)},1}, \sigma^2_{\alpha_k^{(c,u)},1}) + (1 - s_{cu}) \sum_{k=1}^{K-1} \log\mathcal{N}(\alpha_k^{(c,u)}; 0, \tau_{\ell_u} w_u) + s_{cu}\epsilon_{cu} + \text{const},$$

where $\mu_{\alpha_k^{(c,u)},1} = D_k^{(c,u)}/C_k^{(c,u)}$, $\sigma^2_{\alpha_k^{(c,u)},1} = 1/C_k^{(c,u)}$, $k \in [K-1]$. In particular,

$$C_k^{(c,u)} = \frac{1}{\tau_{\ell_u} w_u} + 2 \sum_{g\in d(u)\cap[G]} \sum_{i:Y_i=c, D_i=g} \sum_{m=k}^{K} r_{im} g(\phi_k^{(c,g)}) + \mathbf{1}\{0 \in d(u)\} \left[ 2 \sum_{i:D_i=0} e_{ic} \sum_{m=k}^{K} r_{im} g(\phi_k^{(c,0)}) \right], \tag{A3}$$

$$D_k^{(c,u)} = \sum_{g\in d(u)\cap[G]} \sum_{i:Y_i=c, D_i=g} \left[ \frac{1}{2} r_{ik} - \sum_{m=k+1}^{K} \frac{1}{2} r_{im} - 2 \left( \sum_{m=k}^{K} r_{im} g(\phi_k^{(c,g)}) \sum_{w\in a(g)\setminus\{u\}} E_{q_t}\{s_{cw}\alpha_k^{(c,w)}\} \right) \right]$$
$$+ \mathbf{1}\{0 \in d(u)\} \sum_{i:D_i=0} e_{ic} \left[ \frac{1}{2} r_{ik} - \sum_{m=k+1}^{K} \frac{1}{2} r_{im} - 2 \left( \sum_{m=k}^{K} r_{im} g(\phi_k^{(c,0)}) \sum_{w\in a(0)\setminus\{u\}} E_{q_t}\{s_{cw}\alpha_k^{(c,w)}\} \right) \right] \tag{A4}$$

$$\epsilon_{cu} = E_{q_t} \log\frac{\rho_{c\ell_u}}{1 - \rho_{c\ell_u}} + \sum_{k=1}^{K-1} \frac{\left\{D_k^{(c,u)}\right\}^2}{2C_k^{(c,u)}} - \frac{1}{2} \sum_{k=1}^{K-1} \left[ \log(\tau_{\ell_u} w_u) + \log(C_k^{(c,u)}) \right]. \tag{A5}$$

It is readily seen $q(s_{cu}, \boldsymbol{\alpha}^{(c,u)})$ is jointly a two-component Gaussian mixture with distinct means and variances. In particular, $q(s_{cu})$ is Bernoulli with success probability $p_{cu} = \sigma(\epsilon_{cu})$; conditional on $s_{cu}$, $q(\boldsymbol{\alpha}^{(c,u)} \mid s_{cu})$ is independent Gaussians with means and variances determined by $s_{cu}$ being 1 or 0.

*Step 1e.* Update $q_{t+1}(\boldsymbol{\gamma}^{(u)})$ for each node $u \in \mathcal{V}^*$ of the tree $\mathcal{T}^*$ over $C$ causes by

$$\log q_{t+1}(\boldsymbol{\gamma}^{(u)}) = \mathbb{E}_{q_t(-\boldsymbol{\gamma}^{(u)})} \log H + \text{const} = \sum_{j,k} \log \mathcal{N}(\gamma_{jk}^{(u)}; \mu_{\gamma_{jk}^{(u)},1}, \sigma^2_{\gamma_{jk}^{(u)},1}) + \text{const}, \tag{A6}$$

where $\mu_{\gamma_{jk}^{(u)},1} = B_{jk}^{(u)}/A_{jk}^{(u)}$ and $\sigma^2_{\gamma_{jk}^{(u)},1} = 1/A_{jk}^{(u)}$, $j \in [J]$, $k \in [K]$. In particular,

$$A_{jk}^{(u)} = \frac{1}{\tau^*_{\ell_u^*} w_u^*} + 2 \sum_{c \in d(u) \cap \mathcal{C}} g(\psi_{jk}^{(c)}) \left( \sum_{g=1}^{G} \sum_{i: Y_i = c, D_i = g} r_{ik} + \sum_{i: D_i = 0} e_{ic} r_{ik} \right), \tag{A7}$$

$$B_{jk}^{(u)} = \sum_{c \in d(u) \cap \mathcal{C}} \sum_{g=1}^{G} \sum_{i \in \{Y_i = c, D_i = g\} \cap \mathcal{I}_j} \left\{ r_{ik} X_{ij}^*/2 - 2 r_{ik} g(\psi_{jk}^{(c)}) \sum_{w \in a(c) \setminus \{u\}} E_{q_t}\{s_w^* \gamma_{jk}^{(w)}\} \right\} \tag{A8}$$

$$+ \sum_{c \in d(u) \cap \mathcal{C}} \sum_{i: \{D_i = 0\} \cap \mathcal{I}_j} e_{ic} \left\{ r_{ik} X_{ij}^*/2 - 2 r_{ik} g(\psi_{jk}^{(c)}) \sum_{w \in a(c) \setminus \{u\}} E_{q_t}\{s_w^* \gamma_{jk}^{(w)}\} \right\} \tag{A9}$$

Again it is readily seen that $q_t(\boldsymbol{\gamma}^{(u)})$ is independent Gaussians.

*Step 1f.* Update

$$q_{t+1}(\rho_{c\ell}) = \mathsf{Beta}(a'_{c\ell}, b'_{c\ell}), c \in [C], \ell \in [L],$$

where $a'_{c\ell} = \sum_{u \in \mathcal{V}: \ell_u = \ell} E_{q_t}(s_{cu}) + a_{c\ell}$ and $b'_{c\ell} = \sum_{u \in \mathcal{V}: \ell_u = \ell} \{1 - E_{q_t}(s_{cu})\} + b_{c\ell}$;

For every $d$ steps above, do Step 2-4:

*Step 2.* Update local variational parameters $\boldsymbol{\psi}$ and $\boldsymbol{\phi}$.

$$\phi_k^{(c,g)} = \sqrt{E_{q_t} \left\{ \eta_k^{(c,g)} \right\}^2}, \psi_{jk}^{(c)} = \sqrt{E_{q_t} \left\{ \beta_{jk}^{(c)} \right\}^2}, \tag{A10}$$

for $c \in [C]$, $g \in \{0\} \sqcup [G]$.

*Step 3.* Update the hyperparameters $\boldsymbol{\tau}$ and $\boldsymbol{\tau}^*$.

$$\tau_\ell = \frac{1}{C(K-1) \sum_{u \in \mathcal{V}: \ell_u = \ell} 1} \sum_{u \in \mathcal{V}: \ell_u = \ell} \sum_{c=1}^{C} \sum_{k=1}^{K-1} E_{q_t} \left\{ \left\{ \alpha_k^{(c,u)} \right\}^2 / w_u \right\}, \ell \in [L], \tag{A11}$$

$$\tau^*_\ell = \frac{1}{JK \sum_{u \in \mathcal{V}^*: \ell_u^* = \ell} 1} \sum_{u \in \mathcal{V}^*: \ell_u^* = \ell} \sum_{j=1}^{J} \sum_{k=1}^{K} E_{q_t} \left\{ \left\{ \gamma_{jk}^{(u)} \right\}^2 / w_u^* \right\}, \ell \in [L^*]. \tag{A12}$$

*Step 4.* Compute $\mathcal{E}^*(q_{t+1})$ according to Appendix Appendix C. Stop the iteration once the absolute change in $\mathcal{E}^*(q_{t+1})$ is less than a tolerance `tol=1e-8`. The hyperparameter updates are often slower than the variational parameter updates to converge in terms of the $\mathcal{E}^*(q_{t+1})$. In practice, we can separate the tolerance levels for the hyperparameter updates (`hyper_tol=1e-4`) and VI parameter updates (e.g., `tol=1e-8`). One may update the hyperparameters every $d$ steps of the updates of the variational parameters. In practice, we can adjust $d$ to speed up the convergence. In this paper, we use $d = 10$ which works well in simulations and data analysis. We also suggest multiple initializations to obtain a highest $\mathcal{E}^*(q_{t+1})$ and optimal variational parameters.

#### Appendix B. Calculation of $\log H$

Here we provide the logarithm of the lower bound $H$ for $\mathrm{pr}(\mathcal{D}, \mathbf{\Gamma})$ in Equation (4.20) in the Main Paper.

$$\log H = \sum_{g=0}^{G}\sum_{c=1}^{C}\sum_{i=1}^{N} \mathbf{1}\{Y_i = c, D_i = g\} \Bigg( \log \pi_c^{(g)} \tag{A13}$$

$$+ \sum_{k=1}^{K} \mathbf{1}\{Z_i = k\} \Bigg\{ \sum_{m<k} \Bigg( \log \sigma(\phi_m^{(c,g)}) + (-\eta_m^{(c,g)} - \phi_m^{(c,g)})/2 - g(\phi_m^{(c,g)}) \left\{ \left\{\eta_m^{(c,g)}\right\}^2 - \left\{\phi_m^{(c,g)}\right\}^2 \right\} \Bigg) \tag{A14}$$

$$+ \mathbf{1}\{k < K\} \Bigg( \log \sigma(\phi_k^{(c,g)}) + (\eta_k^{(c,g)} - \phi_k^{(c,g)})/2 - g(\phi_k^{(c,g)}) \left\{ \left\{\eta_k^{(c,g)}\right\}^2 - \left\{\phi_k^{(c,g)}\right\}^2 \right\} \Bigg) \tag{A15}$$

$$+ \sum_{j\in\mathcal{J}_i} \log \sigma(\psi_{jk}^{(c)}) + (X_{ij}^* \beta_{jk}^{(c)} - \psi_{jk}^{(c)})/2 - g(\psi_{jk}^{(c)}) \left\{ \left\{\beta_{jk}^{(c)}\right\}^2 - \left\{\psi_{jk}^{(c)}\right\}^2 \right\} \Bigg\} \Bigg) \tag{A16}$$

$$+ \sum_{c=1}^{C}\sum_{u\in\mathcal{V}}\sum_{k=1}^{K-1} -\frac{1}{2}\log(2\pi\tau_{\ell_u} w_u) - \frac{1}{2\tau_{\ell_u} w_u} \left\{\alpha_k^{(c,u)}\right\}^2 \tag{A17}$$

$$+ \sum_{u\in\mathcal{V}^*}\sum_{j=1}^{J}\sum_{k=1}^{K} -\frac{1}{2}\log(2\pi\tau_{\ell_u^*}^* w_u^*) - \frac{1}{2\tau_{\ell_u^*}^* w_u^*} \left\{\gamma_{jk}^{(u)}\right\}^2 \tag{A18}$$

$$+ \sum_{c=1}^{C}\sum_{u\in\mathcal{V}} [s_{cu}\log\rho_{c\ell_u} + (1 - s_{cu})\log(1 - \rho_{c\ell_u})] \tag{A19}$$

$$+ \sum_{c=1}^{C}\sum_{\ell=1}^{L} [(a_{c\ell} - 1)\log\rho_{c\ell} + (b_{c\ell} - 1)\log(1 - \rho_{c\ell}) - \log \mathsf{B}(a_{c\ell}, b_{c\ell})] \tag{A20}$$

$$+ \sum_{g=0}^{G}\sum_{c=1}^{C} (d_c^{(g)} - 1)\log\pi_c^{(g)} + \text{const}, \tag{A21}$$

where const is a term that does not depend on $\mathbf{\Gamma}$.

#### Appendix C. Calculation of $\mathcal{E}^*(q)$

For ease of presentation, we omit the iterator $t$ in the following. We have $\mathcal{E}^*(q) = E_q \log(H) - E_q \log q + \text{const}$, where the two non-constant terms are:

$$E_q \log(H) = \sum_{g=0}^{G}\sum_{c=1}^{C}\sum_{i=1}^{N} e_{ic}\left\{E_q[\log \pi_c^{(g)}] + \sum_{k=1}^{K} r_{ik} F_{ik}^{(c,g)}(q)\right\} \tag{A22}$$

$$+\sum_{c=1}^{C}\sum_{u\in\mathcal{V}}\sum_{k=1}^{K-1} -\frac{1}{2}\log(2\pi\tau_{\ell_u} w_u) - \frac{1}{2\tau_{\ell_u} w_u} E_q\left\{\alpha_k^{(c,u)}\right\}^2 \tag{A23}$$

$$+\sum_{u\in\mathcal{V}^*}\sum_{j=1}^{J}\sum_{k=1}^{K} -\frac{1}{2}\log(2\pi\tau^*_{\ell^*_u} w^*_u) - \frac{1}{2\tau^*_{\ell^*_u} w^*_u} E_q\left\{\gamma_{jk}^{(u)}\right\}^2 \tag{A24}$$

$$+\sum_{c=1}^{C}\sum_{u\in\mathcal{V}} E_q\{s_{cu}\} E_q \log \rho_{c\ell_u} + (1 - E_q\{s_{cu}\}) E_q \log(1-\rho_{c\ell_u}) \tag{A25}$$

$$+\sum_{c=1}^{C}\sum_{\ell=1}^{L} (a_{c\ell}-1) E_q \log \rho_{c\ell} + (b_{c\ell}-1) E_q \log(1-\rho_{c\ell}) - \log \mathsf{Beta}(a_{c\ell}, b_{c\ell}) \tag{A26}$$

$$+\sum_{g=0}^{G}\sum_{c=1}^{C} (d_c^{(g)}-1) E_q \log \pi_c^{(g)} - \sum_{g=0}^{G} \log \mathsf{B}(\boldsymbol{d}^{(g)}), \tag{A27}$$

where $\mathsf{B}(\boldsymbol{x} = (x_1, \ldots, x_I)) = \prod_i \Gamma(x_i)/\Gamma(\sum_i x_i)$ and $\Gamma(\cdot)$ is the Gamma function, $x_i > 0, i \in [I]$ (when $I = 2$, $B(\cdot)$ is the Beta function); and

$$-E_q \log q = -\sum_{g=0}^{G}\left(\sum_{c=1}^{C}\left(\sum_{i=1}^{N} e_{ic} + d^{(c,g)} - 1\right) E_q\{\log(\pi_c^{(\cdot,g)})\} - \log \mathsf{B}(\sum_{i=1}^{N} e_{ic} + d^{(c,g)}, c = 1, \ldots, C)\right) \tag{A28}$$

$$+0.5\sum_{c=1}^{C}\sum_{u\in\mathcal{V}}\sum_{k=1}^{K-1} E_q\{s_{cu}\} + E_q\{s_{cu}\}\log(2\pi\sigma^2_{\alpha_k^{(c,u)},1}) \tag{A29}$$

$$+0.5\sum_{c=1}^{C}\sum_{u\in\mathcal{V}}\sum_{k=1}^{K-1} E_q\{1-s_{cu}\} + E_q\{1-s_{cu}\}\log(2\pi\tau_{\ell_u} w_u) \tag{A30}$$

$$-\sum_{i:D_i=0}\sum_{c=1}^{C} e_{ic}\log e_{ic} - \sum_{i=1}^{N}\sum_{k=1}^{K} r_{ik}\log r_{ik} \tag{A31}$$

$$-\sum_{c=1}^{C}\sum_{u\in\mathcal{V}} \{E_q[s_{cu}]\log(p_{cu}) + E_q[1-s_{cu}]\log(1-p_{cu})\}$$

$$-\sum_{c=1}^{C}\sum_{\ell=1}^{L}\left\{(a'_{c\ell}-1) E_q\{\log\rho_{c\ell}\} + (b'_{c\ell}-1) E_q\{\log(1-\rho_{c\ell})\} - \log \mathsf{B}(a'_{c\ell}, b'_{c\ell})\right\} \tag{A32}$$

#### Appendix D. Additional Details of Simulation Studies

*Simulation I* We use the domain hierarchy with $p_{\mathsf{leaf}} = 6$ domain leaves and 2 non-root nodes with root node $u = u_0 = 1$ (see Figure 2(a) in the Main paper). The total number of causes is $C = 3$. We set the total sample sizes to be $N = 1000$ with the domain-specific sample sizes being 1) evenly and randomly distributed across domains or 2) unevenly and randomly allocated by domain: we first form pairs of domains and evenly and randomly allocated the total sample sizes to all the pairs of domains; then within each pair, we randomly allocate samples with a ratio of 4 to 1. In addition, we set $G = 5$ source domains and 1 target domain; the number of latent classes for each cause is $K = 2$, for $J = 20, 60$ binary responses. We considered two scenarios of the response probability profiles: 1) stronger signal: $\theta_{j1}^{(c,g)} = 0.95$, $\theta_{j2}^{(c,g)} = 0.05$; 2) weaker signal: $\theta_{j1}^{(c,g)} = 0.8$, $\theta_{j2}^{(c,g)} = 0.2$.

Two scenarios of between-domain patterns of CSMFs are considered: 1) balanced: $\pi_c^{(g)} = 1/C$, and 2) unbalanced: $\boldsymbol{v}^{(g)} = (x_1, x_2, \ldots, x_C)/C$ and $x_c = 5$ if $c = 1$, and $x_c = 3$ if $c \not\equiv 0 \pmod{C}$, $c = 1, \ldots C$. We picked the target domain CSMF to be $\boldsymbol{\pi}^{(0)} = \boldsymbol{v}^{(3)}$ and $\boldsymbol{\pi}^{(g)}$, $g = 1, \ldots, G$ to take the rest of vectors: $\boldsymbol{v}^{(1)}, \boldsymbol{v}^{(2)}, \boldsymbol{v}^{(4)}, \ldots, \boldsymbol{v}^{(G)}$.

For each domain, the class mixing weights $\boldsymbol{\lambda}^{(c,g)}$ are generated independently for each cause $c$ by the following scheme: 1) for cause $c$, sample independently $\boldsymbol{\alpha}^{(c,u_0)}$ for the root domain node: $\boldsymbol{\alpha}^{(c,u_0)} \sim \boldsymbol{F}(\mathsf{Dirichlet}(2, K))$, where $\boldsymbol{F}$: $\mathcal{S}^{K-1} \to \mathbb{R}^{K-1}$ maps a vector in the $K$-probability simplex to a vector in the $K-1$ dimensional Euclidean space $\boldsymbol{F}(\boldsymbol{\lambda}) = \boldsymbol{\alpha}$ where $\boldsymbol{\alpha} = (\alpha_1, \ldots, \alpha_{K-1})$ is the unique vector that satisfies $\lambda_1 = \sigma(\alpha_1)$, …, $\lambda_k = \sigma(\alpha_k)\prod_{s<k}(1-\sigma(\alpha_s))$, …, and $\lambda_K = \prod_{s<K}(1-\sigma(\alpha_s))$; 2) For cause $c$, set the same and fixed diffusions upon $\boldsymbol{\alpha}^{(c,u)}$ for non-root nodes $u$ to be $-2$ if $u = 2$, 2 if $u = 3$, and zero for $u \geqslant 3$.

The simulation setup creates a scenario the True Domain Grouping of four blocks: {0, 1}, and {2, 3}, {4}, {5}, . The Complete Pooling approach sets $s_{cu} = 0$ for any non-root node $u \in \mathcal{V} \setminus u_0$, forming a single group of six domains. The Ad Hoc Domain Grouping method splits {0, 1} into {0} and {1} resulting in a finer domain grouping. For the No Domain Grouping approach, we share the class-specific response profiles, but do not borrow information across domains to perform shrinkage about the mixing weights $\boldsymbol{\lambda}^{(c,g)}$, $g \in \{0\} \cup [G]$. In the method Domain Adaptive, we used hyperparameters $a_{c\ell} = b_{c\ell} = 1$ in the selection probability of the spike-and-slab prior along the domain hierarchy. For all approaches, we set $\boldsymbol{d}^{(g)} = (1, \ldots, 1)$ for all the domains. During estimation, we use a two-level cause tree with a root node pointing towards $C$ cause leaves with equal edge weights.

*Appendix D.1 Simulation IIa and IIb*

*Design* Two designs (referred to as IIa and IIb) are considered where the difference lies in how the masked CODs are chosen, in a uniform or non-uniform fashion over the causes.

In Simulation IIa), we randomly split subjects into 80% training and 20% testing data. We then collect the 20% split from each PHMRC site into a single target domain on which CSMF and CODs are to be inferred. In this basic setup, the causes-of-deaths in the target domain are close to the population average across domains; the conditional distributions of the VA responses given the cause is also close to the counterpart estimates based on data from the source domains.

In Simulation IIb) for each cause, we draw a random fraction of deaths $\varphi_c$ generated from a half-half mixture: $0.5\mathsf{Beta}(1,5) + 0.5\mathsf{Beta}(1,20)$; $\varphi_c$ is also independently generated across causes, so that when constructing the target domain data some causes are up-sampled while others are down-sampled relative to the global CSMFs. We have designed such a scheme to let the constructed target domain to have a CSMF that is different from those in the source domains. We then collect the sampled deaths into a single domain, and treat it as the target domain on which the CSMFs and CODs are to be inferred. In this setup, the target may have distinct CSMFs from other domains; the conditional distribution of VA responses given a cause is a mixture across the other domains. In both cases, the domain trees have $p_{\mathsf{leaf}} = 7$ leaves and $p - p_{\mathsf{leaf}} = 3$ non-leaf nodes. Note that because the constructed target domain is a random sample from the entire data, we specify weights for the edges in the tree so that the tree-based distance from the constructed target domain to the six original domains are identical.

*Results* Figure Appendix Figure 1 shows the relative comparisons of the various options of conducting target domain CSMF estimation in terms of CMSF accuracy; unlike Simulation I, here the True Domain Grouping comparator is unavailable. In particular, the domain adaptive method which adaptively encourage shrinkage along the domain hierarchy produced estimates with slightly better accuracy overall. In addition, the task of CSMF estimation in the constructed target domain is more challenging when CSMFs differ substantially from the source domains.

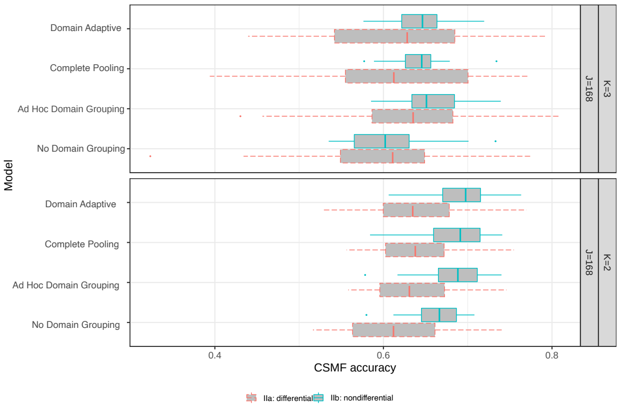


(a) CSMF Accuracy Comparison

Appendix Figure 1: Simulation IIa and IIb show the proposed method achieves better estimation accuracy in terms of CSMF accuracy.

#### Appendix E. Tree-Structured Shrinkage Priors: A Review

We specify a prior distribution for a set of real-valued parameters without range constraints that may differ by leaf nodes $\{\vartheta_v : v \in \mathcal{V}_{\mathsf{leaf}}\}$. In specifying the tree-structure shrinkage prior, we need a few pieces of tree-related information: a weighted rooted tree $\mathcal{T}_w = (\mathcal{T} = (\mathcal{V}, E), w)$ with leaves $\mathcal{V}_{\mathsf{leaf}} \subset \mathcal{V}$, edge lengths $\boldsymbol{w} = (w_u)_{u \in \mathcal{V}}$, the leaf id for each observation $\mathcal{L} = (v_1, \ldots, v_N)^\mathsf{T}$ where the sample-to-leaf indicator $v_i$ chooses parameter $\vartheta_{v_i}$ to partly characterize the distribution of data from subject $i$. Because leaf-specific sample sizes may vary, we propose a tree-structured prior to borrow information across nearby leaves. The prior encourages collapsing certain parts of the tree so that observations within a collapsed leaf group share the same parameter value. Li *and others* (2021) has extended Thomas *and others* (2020) to deal with rooted weighted trees.

We specify a spike-and-slab Gaussian diffusion process prior along a rooted weighted tree for $\vartheta_v$. For a leaf $v \in \mathcal{V}_{\mathsf{leaf}}$, let

$$\vartheta_v = \sum_{u \in a(v)} \varphi_u. \tag{A33}$$

Here $\vartheta_v$ is defined for leaves only and $\varphi_u$ is defined for all the nodes. Suppose $v$ and $v'$ are leaves and siblings in the tree such that $pa(v) = pa(v')$, setting $\varphi_v = \varphi_{v'} = 0$ implies $\vartheta_v = \vartheta_{v'}$. More generally, a sufficient condition for $M$ leaves $\vartheta_v$, $v \in \{v_1, \ldots, v_M\}$ to fuse is to set $\varphi_u = 0$ for any $u$ that is an ancestor of any of $\{v_1, \ldots, v_M\}$ but not common ancestors for all $v_m$. That is, to achieve grouping of observations that share the same vector of latent class proportions, in our model, it is equivalent to parameter fusing. In the following, we specify a prior on the $\varphi_u$ that *a priori* encourages sparsity, so that closely related observations are likely grouped to have the same vector of class proportions. The fewer distinct ancestors two nodes have, the more likely the parameters $\vartheta_v$ are fused, because the prior would encourage fewer auxiliary variables $\varphi_u$ to be set to zero. In particular, we specify

$$\varphi_u = s_u \alpha_u, \forall\ u \in \mathcal{V}, \tag{A34}$$

$$\alpha_u \sim N(0, \tau_{\ell_u} w_u),\ \text{independently for } \forall\ u \in \mathcal{V}, \tag{A35}$$

$$s_{u_0} = 1,\ \text{and } s_u \sim \mathsf{Bernoulli}(\varrho_{\ell_u}),\ \text{independently for } u \in \mathcal{V} \setminus u_0, \tag{A36}$$

$$\varrho_\ell \sim \mathsf{Beta}(a_\ell, b_\ell),\ \text{independently for } \ell \in [L], \tag{A37}$$

where $N(m, s)$ represents a Gaussian density function with mean $m$ and variance $s$. $\tau_\ell$ is the unit-length variance and controls the degree of diffusion along the tree which may differ by node level $\ell_u$ where $\ell_u \in [L]$ represents the "level" or "hyperparameter set indicator" for node $u$. For example, in simulations and data analysis, we will assume that the root for the diffusion process has a prior unit-length variance distinct from other non-root nodes. For the root $u_0$ with $s_{u_0} = 1$, $\alpha_{u_0}$ initializes the diffusion of $\vartheta_u$.

Leaf groups are formed by selecting a subset of nodes in $\mathcal{V}$: $\mathcal{U} = \{u \in \mathcal{V} : s_u = 1\}$. Except a probability-zero set, two leaves $v$ and $v'$ are grouped, or "fused", if and only if $a(v) \cap \mathcal{U} = a(v') \cap \mathcal{U}$. In particular, the null set is $\{\vartheta_v = \vartheta_{v'}\} \cap \{\sum_{u \in [a(v)\cap\mathcal{U}]\setminus[a(v')\cap\mathcal{U}]} \alpha_u = \sum_{u \in [a(v')\cap\mathcal{U}]\setminus[a(v)\cap\mathcal{U}]} \alpha_u\}$ where the latter has probability zero. We may estimate $\mathcal{U}$, e.g., using the posterior median model.

REMARK Appendix E.1 Equations (A33)-(A37) define a Gaussian diffusion process initiated at $\alpha_{u_0}$:

$$\vartheta_u \mid \{\varphi_{u'}, u \in a(u)\}, s_u, \tau_{\ell_u}, w_u \sim N\left(\sum_{u' \in a(u)} \xi_{u'}, s_u \tau_{\ell_u} w_u\right), \tag{A38}$$

for any non-root node $u \neq u_0$; also see the seminal formulation by Felsenstein (1985). To aid the understanding of this Gaussian diffusion prior, it is helpful to consider a special case of $s_u = 1$ and $\ell_u = 1$, $\forall u \in \mathcal{V}$. For two leaves $v, v' \in \mathcal{V}_{\mathsf{leaf}}$, the prior correlation between $\vartheta_v$ and $\vartheta_{v'}$ is

$$\text{Corr}(\vartheta_v, \vartheta_{v'}) = \frac{\sum_{u \in a(v)\cap a(v')} w_u}{\{dist_{\mathcal{T}_w}(u_0, v) dist_{\mathcal{T}_w}(u_0, v')\}^{1/2}}, \tag{A39}$$

When $v$ and $v'$ have the same number of ancestors ($|a(v)| = |a(v')|$) and all edges have identical weight $w_u = c, \forall u$, the prior correlation is the fraction of common ancestors.

### Appendix F. Appendix Figures

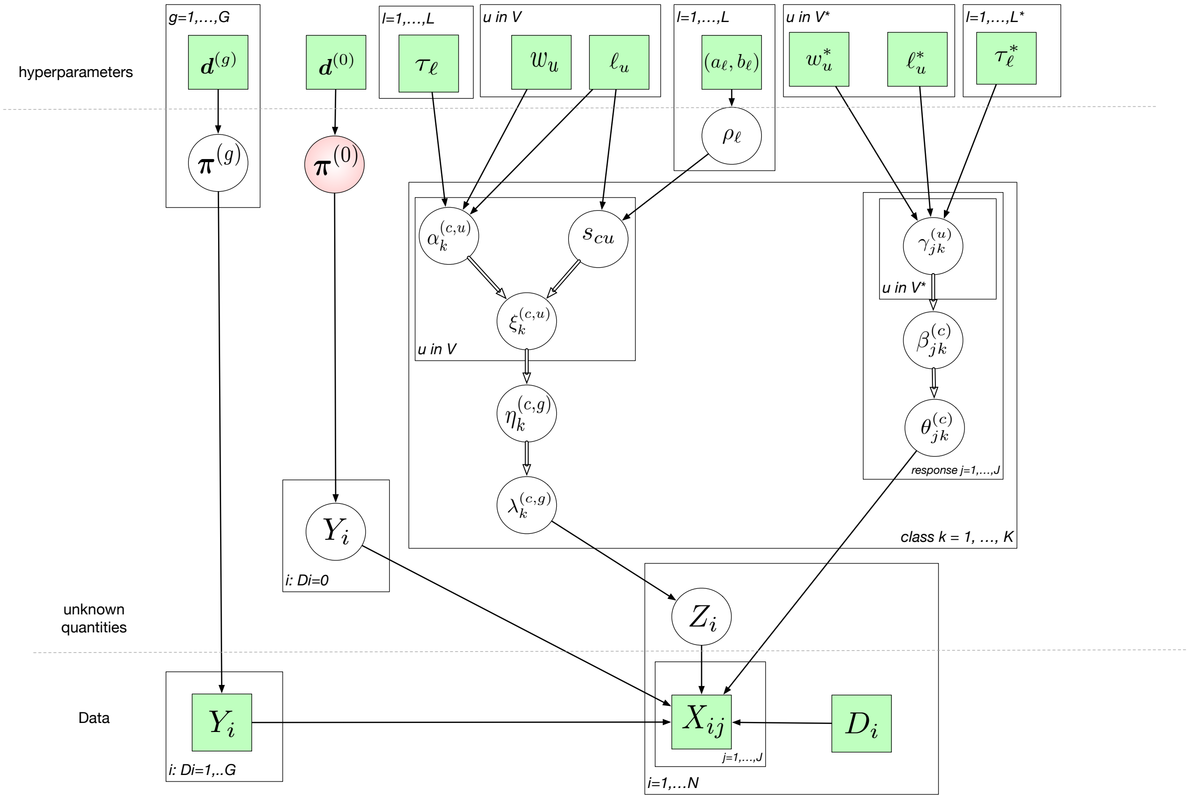


Appendix Figure 2: The directed acyclic graph (DAG) representing the structure of the model likelihood and priors following the style of Koller and Friedman (2009). The quantities in squares are either data or hyperparameters; the unknown quantities are shown in the circles; the double-stroke circle $Z_i$ indicates a selector, choosing the latent class $k = 1, \ldots, K$. The arrows connecting variables indicate that the parent parameterizes the distribution of the child node (solid lines) or completely determines the value of the child node (double-stroke arrows). The rectangular "plates" where the variables are enclosed indicate that a similar graphical structure is repeated over the index; The index in a plate indicates nodes, hyperparameter levels, leaves, subjects, classes and features. The parameter of interest $\boldsymbol{\pi}^{(0)}$, the CSMFs in the target domain, is highlighted.

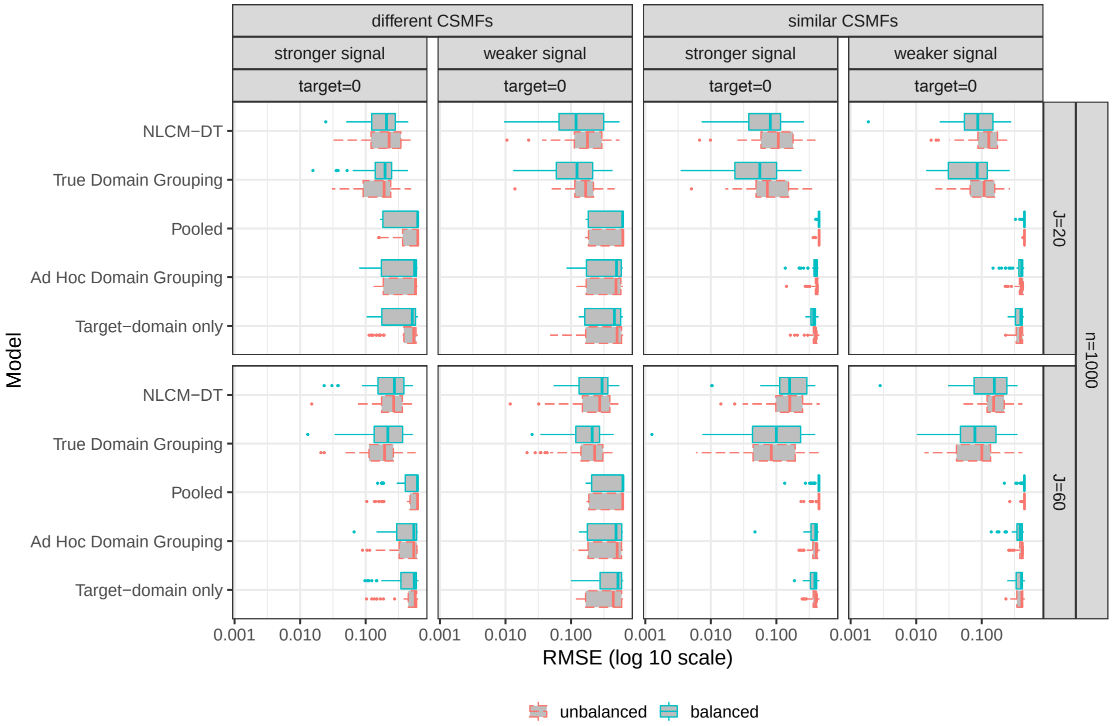


Appendix Figure 3: Simulation I: RMSE comparison.
